# Urinary extracellular vesicles for high-precision bladder cancer subtyping and prognosis

**DOI:** 10.1101/2025.10.14.25338041

**Authors:** Zachary F. Greenberg, Tarun E. Hutchinson, Johnathan Kahn, Kiley S. Graim, Padraic O’Malley, Mei He

**Author notes:** Corresponding Authors and Padraic.O’.

## Abstract

Bladder cancer exhibits molecular heterogeneity that complicates early diagnosis and prognosis, and drives confounding clinical outcomes. Non–muscle invasive and muscle-invasive subtypes, especially for intermediate to high grade, carry a 25 – 50% progression-free survival rate, underscoring the need for high precision prognostic strategy. Urinary extracellular vesicles (uEVs) are promising carriers of tumor-derived RNAs and proteins. However, significant challenges in studying uEVs arise from the diverse cellular origin of uEVs associated with the dynamic composition of urine, which presents roadblocks for developing the clinical utility of uEVs. We introduced an AI-driven EV liquid biopsy pipeline that integrates (1) standardized EV isolation via NanoPom magnetic beads, (2) transcriptomic profiling for molecular subtyping, and (3) prognostic scoring algorithm. In a discovery cohort of 16 bladder cancer patients including both MIBC and NMIBC, we compared NanoPom isolated uEVs with ExoEasy and Fujifilm MagCapture isolated uEVs, for identifying bladder cancer subtype-specific gene signatures, and externally validated them using UCSC Xena. The result outperformed currently reported bladder cancer diagnostic biomarkers from assays including Galeas, CxBladder, and Xpert. In a validation cohort of matched 7 patient plasma samples, we confirmed with plasma derived EVs for correlating with urinary EV biomarkers from NGS sequencing. The prognostic score stratified patients into low-, intermediate-, and high-grade risk groups based on Xena’s bladder cancer survival outcomes. Our AI-driven uEV liquid biopsy pipeline proves the concept for high precision bladder cancer subtyping and prognosis, which could potentially facilitate treatment decision and lead to advanced profiling of bladder tumor biology using uEV liquid biopsy.

## Introduction

Urothelial bladder cancer (BCa) is one of the top 10 common cancers globally. The muscle-invasive bladder cancer (MIBC) is the most aggressive subtype with residual tumor or node-positive disease, leading to only a 25% 5-year survival rate after treatment of radical cystectomy and pelvic lymph node dissection (RC-PLND)^1, 2^. The alternative (non-surgical neoadjuvant chemotherapy (NAC)) is underutilized to overcome the complication and morbidity from RC-PLND, largely due to the lack of a non-invasive prognostic and predictive strategy for MIBC. The molecular mechanisms of invasive MIBC phenotypes and their progression are poorly understood. Currently, MIBC is primarily characterized by distinct molecular pathways, including specific genomic alterations that differ from those in non-muscle-invasive bladder cancer (NMIBC)^3^. Specifically, MIBC tumor-derived extracellular vesicles (EVs) have been extensively reported to regulate molecular interactions between tumor cells and other stromal or immune cells in the tumor microenvironment (TME), thereby altering the TME environment. Such EV-based signaling regulations can mediate physiological and pathophysiological processes in the generation of an invasion-supporting environment and the formation of premetastatic niches^4–6^. Therefore, molecularly mapping EV profiles at the cellular level could offer unmatched opportunities for elucidating the mechanism of MIBC progression and associated therapeutic responses from a liquid biopsy.

Urinary EVs (uEVs) are currently emerging as a precision diagnostic and prognostic approach. However, the diverse cellular origins of uEVs and the dynamic composition of urine^7–9^ often pose substantial challenges for reliable molecular interpretation, which may confound downstream investigations. The use of different EV isolation methods also led to significant variation and non-reproducible biomarker discovery. In order to overcome complexity and heterogeneity, we introduced an AI-driven BCa EV liquid biopsy pipeline, which is composed of our previously developed NanoPom EV isolation strategy^10^ and transcriptomic analyses to identify BCa biomarkers that subtype BCa into MIBC and NMIBC, and deploy a novel EV-based prognostic strategy using our algorithm, ExoMasso, to determine molecular grading in each BCa subtype. Previously, no EV-based models existed that rigorously inferred patient tumor biomarker mRNA signatures using EV-derived mRNA counts as input for the prediction. Therefore, we developed ExoMasso to address this major challenge by leveraging an autoencoder-based deep learning architecture^11^ that uses a maximum mean discrepancy^12^ with non-negative weights as the loss function to predict tumor mRNA signatures in patients from a series of patient-based EV-derived mRNA transcripts. Upstream, ExoMasso utilizes DESeq2-determined differentially abundant transcripts that differentiate MIBC from NMIBC in both EVs and sequenced bladder tissues. These differential transcripts enable ExoMasso to infer how uEV mRNA signatures can be mapped to BCa tumor mRNA signatures using UCSC Xena’s dataset (Xena)^13^. After inference, ExoMasso then leverages a Cox-Hazard Proportional Hazard model (CoxPH)^14^ to molecularly grade BCa subtypes and generate prognostic scores. We trained our CoxPH model on the Xena dataset that was stratified into NMIBC and MIBC using our uEV biomarkers and the associated overall survival data.

We first demonstrated the proof-of-concept in 16 patients with bladder cancer. Following high-quality uEV isolation, we performed transcriptomic analysis to identify BCa biomarkers that subtyped patients into NMIBC and MIBC, which were then externally validated in the Xena dataset. Since uEVs contain many mRNA transcripts, we leveraged our transcriptomic dataset to demonstrate how our discovered mRNA markers differentiate NMIBC and MIBC in Xena’s bladder cancer dataset compared to commercial gene sets from Galeas^15, 16^, CxBladder^17, 18^, and Xpert^19, 20^. These commercial assays sample urine to collect DNA^15, 16^ and mRNAs^17–20^ from urinary cell pellets. However, uEVs may also contribute significantly to these detection tests since they also carry these respective DNA and mRNA markers. Next, we rigorously validated our discovered biomarkers by evaluating whether their mRNA signature in uEVs correlate with plasma-derived EVs. In a validation cohort of matched 7 patients, we demonstrated that both plasma- and urine-derived EVs exhibit highly similar biomarker concordance, supporting the potential of our BCa-based uEV liquid biopsy for early diagnosis and subtyping. Finally, upon deploying ExoMasso, we stratified patients into low-, intermediate-, and high-grade risk groups based on Xena’s survival outcomes. Our AI-driven EV liquid biopsy pipeline (**Figure 1**) enables clinical translation for the early detection and prognosis of BCa subtypes, which leads to advanced profiling of bladder tumor biology based on circulating EVs.

**Figure 1.**
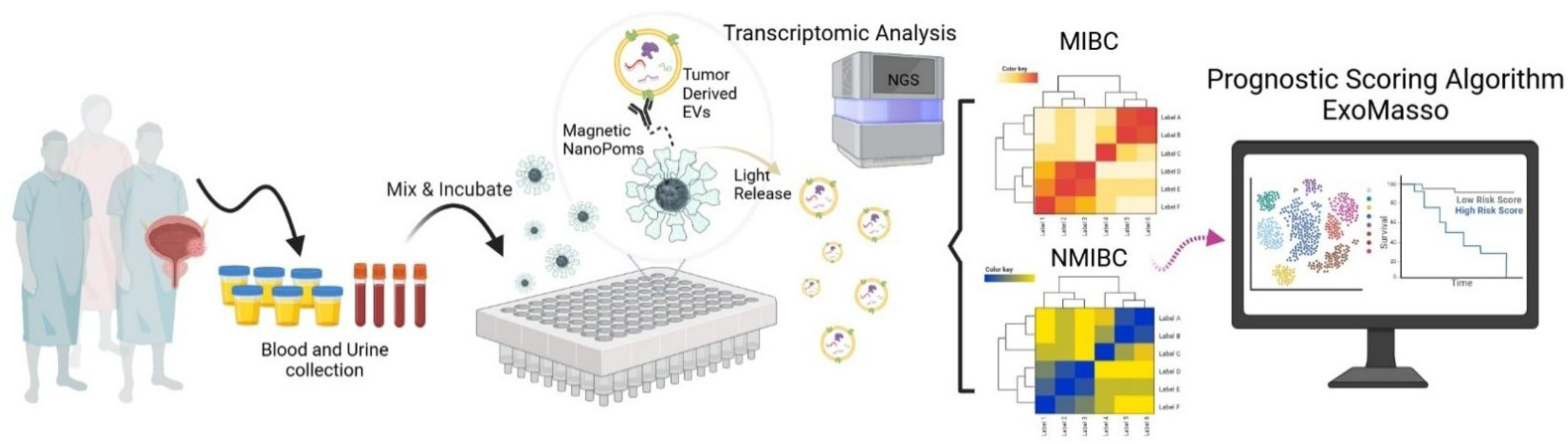
Schematic illustration of an AI-driven EV liquid biopsy pipeline that integrates (1) standardized EV isolation via NanoPom magnetic beads, (2) transcriptomic profiling for molecular subtyping, and (3) prognostic scoring algorithm.

## Materials and Methods

### Urine and Plasma Collection & Sample Preparation

#### NanoPom Fabrication and Isolation

Our previously published unique nanographene pom pom-like microbeads (NanoPom)^10^. Briefly, the NanoPom is fabricated by using a Fe_3_O_4_/SiO_2_ core, followed by a series of sequential layer depositions of nanographene to form pom-like sheet layers with nanocavity in between, which enables a size hindrance to avoid considerably larger membrane structures. Streptavidin is cross-linked onto the beads, followed by biotinylated antibody conjugation (CD9, CD63, and CD81) are added. To capture EVs, 40 μl (approx. 0.4 mg beads) of the NanoPoms was mixed with 1 mL of urine or plasma. Afterward, the sample mixture was incubated on a revolver for 24 hour at 4°C. After incubation, captured EVs were magnetically aggregated to remove the liquid solution for washing 3 times with ice-cold, 1x PBS. Subsequently, the washed beads solution in 40 μl was transferred to a 200 uL solution of ice-cold 1x PBS containing 25 uM trehalose for 20 minute expose to 365 nm^-1^ UV to release captured EVs.

#### ExoEasy Maxi Kit for urinary EV isolation

ExoEasy (QIAGEN) was used according to the manufacturer’s protocol. 1 mL of urine was added to an equal volume of XBP reagent, mixed, and centrifuged at 500x RCF for 5 minutes at room temperature. The membrane was washed with XWP reagent using 3000x RCF for 5 minutes at room temperature, followed by adding 400 uL of XE reagent for a 5-minute centrifugation at 500x RCF at room temperature. We reapplied the eluate to the column, as per manufacturer recommendation, and centrifuged for 3000x RCF for 5 minutes to obtain EVs.

#### Fujifilm MagCapture for urinary EV isolation

Fujifilm’s MagCapture Exosome Isolation Kit PS v1 was used per the manufacturer’s protocol. 60 uL of magnetic beads were combined with 500 uL of Exosome Capture Immobilizing Buffer, mixed, and supernatant removed. 10 uL of Biotin-labeled Exosome Capture reagent was added to the beads in 500 uL of Exosome Capture Immobilizing Buffer for 10 minutes at 4°C. The beads were washed 3x with 500 uL of the Exosome Capture Immobilizing Buffer. Notably, 2 uL of Exosome Binding Enhancer was added to 1 mL of urine sample. Washed beads were magnetically removed, applied to the urine sample, and incubated for over 3 hours at 4°C. After incubation, 6 uL Exosome Binding Enhancer was added to 3 mL of Exosome Washing Buffer. This washing buffer was used to wash EV-captured beads 3x, followed by eluting twice with 50 uL of the Exosome Elution Buffer for a final eluate volume of EVs in 100 uL.

#### Nanoparticle Tracking Analysis

Nanoparticle tracking analysis was conducted using ZetaView (QUATT, Particle Metrix Inc, USA). ZetaView measures the nanoparticle’s Brownian motion using an incident laser to determine the corresponding size. The nanoparticle motion is then tracked by the detector and recorded over time: The incident laser wavelength was 488 nm^-1^ with sensitivity at 75 and shutter time at 163, over 90 seconds at the highest video resolution for all 11 positions.

#### Protein Extraction and BCA Quantification

18 uL of Isolated EVs were lysed with 2 uL of 1x ice-cold RIPA buffer. Samples were incubated on ice for 15 minutes, with 30-second vortexing every 5 minutes. At the end of the incubation, the samples were sonicated for 15 seconds. Total protein was then quantified with the Pierce BCA Protein Assay Kit (Thermo Fisher, USA). Per the vendor’s protocol, reagents A and B were mixed at a 24:1 ratio to formulate the working buffer. In a separate tube, 10 uL of the working reagent and lysed EV sample were added, followed by incubation at 75°C for 5 minutes, then cooled to room temperature to measure 562 nm^-1^ absorbance. Known quantities of bovine serum albumin were utilized to generate the standard calibration curve.

#### ExoView R100

The ExoView R100 was utilized for EV tetraspanin (CD9, CD63, and CD81) abundance levels per standard protocols provided by NanoView Biosciences (Now Unchained Labs). After NTA evaluation of particles isolated by each of the methods, all inputs to the ExoView R100 were standardized to the lowest particle concentration by using solution A (pH 7.4), then all samples were diluted to the manufacturer’s recommendation for particles obtained from plasma. Next, 40 uL of diluted sample was dropped onto the microarray chip, incubated overnight, and the assay was conducted as per manufacturer’s instructions.

#### Transcriptomic Analysis

Transcriptomic analysis was conducted through a modified FREYA pipeline^21^. First, read quality control and trimming was performed by fastp^22^, followed by read mapping to hg38 and quantifying mapped reads with Salmon^23^. Next, Tximport^24^ was used to aggregate mapped reads into a count matrix. Batch correction was performed using ComBat-seq^25^. Using DESeq2^26^, transcripts were filtered with counts <= 10 with at least 3 samples having these counts and differential expression testing was performed. DESeq2 uses a negative binomial distribution for the generalized linear model explaining bladder cancer subtyped gene expression differences while adjusting for each sample’s contribution to the observed EV isolate mRNA. For visualizations, samples were transformed using DESeq2’s variance stabilizing transformation. Transcripts passing the false discovery rate cutoff were used in downstream enrichment analysis and comparison to Xena. Enrichment analysis of statistically significant transcripts was performed using gProfiler^27^ and the HumanBase^28^ community detection algorithm to identify tissue-specific functional network interactions enriched with differentially expressed transcripts. HumanBase builds genome-scale functional map of human tissues to serve as a profiler of gene-specific function within tissue networks. HumanBase will attempt to profile input genes with any interacting partners, illustrated as the interaction confidence. Supervised UMAP and LDA were performed to investigate the molecular space for urinary and plasma EV signatures. Our UMAP parameters are listed in Table S3. To evaluate urinary EV mRNA signature concordance with UCSC Xena, a batch correction was performed with all datasets. Next, UCSC Xena was projected into the urinary EV molecular space using our UMAP model, then labelled as MIBC or NMIBC based on the nearest neighbor distance to the closest urinary EV cluster.

#### ExoMasso

ExoMasso is an autoencoder-based deep learning model that utilizes uEV mRNA signatures as input to predict either MIBC or NIBC mRNA signatures as output. The training data was previously batch corrected using ComBat-seq. The encoder-and-decoder networks consist of 2 fully connected layers with Rectified Linear Unit (ReLU)^29^ activations, constrained by non-negative weights with gradient clipping to ensure biologically meaningful interpretations each mRNA contributes to ExoMasso’s training. The objective function we used was the Maximum Mean Discrepancy (MMD) between predicted and true BCa mRNA signatures using a radial basis kernel with the bandwidth tuned by the median pairwise distance. MMD is a measure of non-parametric distance between two distributions in a kernelized embedded space. Since MMD aims to minimize the means, this approach enables meaningful granularity in the predicted BCa mRNA signatures while centralizing onto the known BCa mean mRNA signature. We further constrain ExoMasso to use non-negative weights in the model to ensure meaningful explanations to how each mRNA contributes to the predicted BCa mRNA signature. Adaptive momentum (Adam)^30^ was used to optimize ExoMasso. A multi-seed K-fold cross-validation strategy was used to evaluate ExoMasso’s generalization of the stratified Xena dataset, determined by previous nearest neighbor analysis in our uEV-based UMAP embedded space. ExoMasso’s model training proceeded until the average Wasserstein distance for the mRNA signature was less than 1. Performance was assessed by the grand-average correlation between predicted and true mRNA expression in BCa samples.

#### Cox-Hazard risk model and prognostic scoring for subtyped bladder cancer

To generate a prognostic score from the predicted BCa mRNA signatures, a Cox-Hazard risk model^14^ was composed using Xena’s patient overall survival data and the uEV-based mRNA signature. The Cox-Hazard model will generate risk score based on the molecular subtype and associated survival. We partitioned the risk score into 3 quantiles corresponding to low, intermediate, and high grade tumors. Next, BCa-predicted mRNA signature corresponding to the predicted bladder cancer subtype from our uEV’s mRNA signature is evaluated by the cox-hazard model to generate associated grades and mortality odds for each patient.

#### Statistics

All plots with error bars mean ± standard deviation (N=4 per sample). Analyses and plotting were performed with Python v3.10, R v4.4 and GraphPad v10.0. The false discovery rate for statistically different transcripts was calculated by Benjamini-Hochberg^31^.

## Results

### High-quality uEV isolation by NanoPoms

We benchmarked NanoPoms immunomagnetic isolation^10^ against other commercial EV isolation assays, including MagCapture and ExoEasy, to ensure that high-quality uEVs were obtained before conducting next-generation sequencing. Indicated by previous reports^32–34^, we observed that the uEV heterogeneity is dependent on the isolation assay performed using nanoparticle tracking analysis, showing differences in total EV protein (Figure 2A), EV yield (Figure 2B), protein per EV (Figure 2C), and EV size (Figure 2D) despite being isolated from the same bladder cancer patient samples. The representative particle distribution across the isolation methods was shown in Figure 2E. NanoPoms yields a majority of particles around 100 nm in diameter. In comparison, ExoEasy and MagCapture yielded particles in larger size, ranging from 100 to 200 nm. In order to corelate the quality of purified uEVs with bladder tumor origin, we used flow cytometry NanoAnalyzer (NanoFCM) to characterize the EV particle concentration (Figure 2F), size (Figure 2G), and bladder tumor marker MUC4 expression level (Figure 2H), which revealed that NanoPom isolation method delivered the majority of EV particles in high expression of MUC4, although the total particle concentration is lower than ExoEasy method, indicating the higher relevance of NanoPom isolated EVs to bladder tumor origin. ExoEasy isolation method delivered a different population of EV particles with much fewer expressions of bladder tumor marker MUC4. When we evaluated the well-known bladder cancer hallmark, MUC4, through the ExoView platform^35^, we observed a significant proportion of NanoPom-isolated uEVs presented MUC4 colocalized with EV marker CD63, compared to ExoEasy and MagCapture (Figure 2I). Comparatively, ExoEasy was observed to isolate uEVs with the lowest marker expression. Upon viewing the fluorescent microscopic images, we observed significant debris associated with MagCapture, and few debris associated to ExoEasy, and none by the NanoPoms (Figure 2J). Our results demonstrate that using NanoPoms is essential for obtaining high-quality uEVs from bladder tumor origin for biomarker discovery to differentiate between MIBC and NIBC signatures.

**Figure 2.**
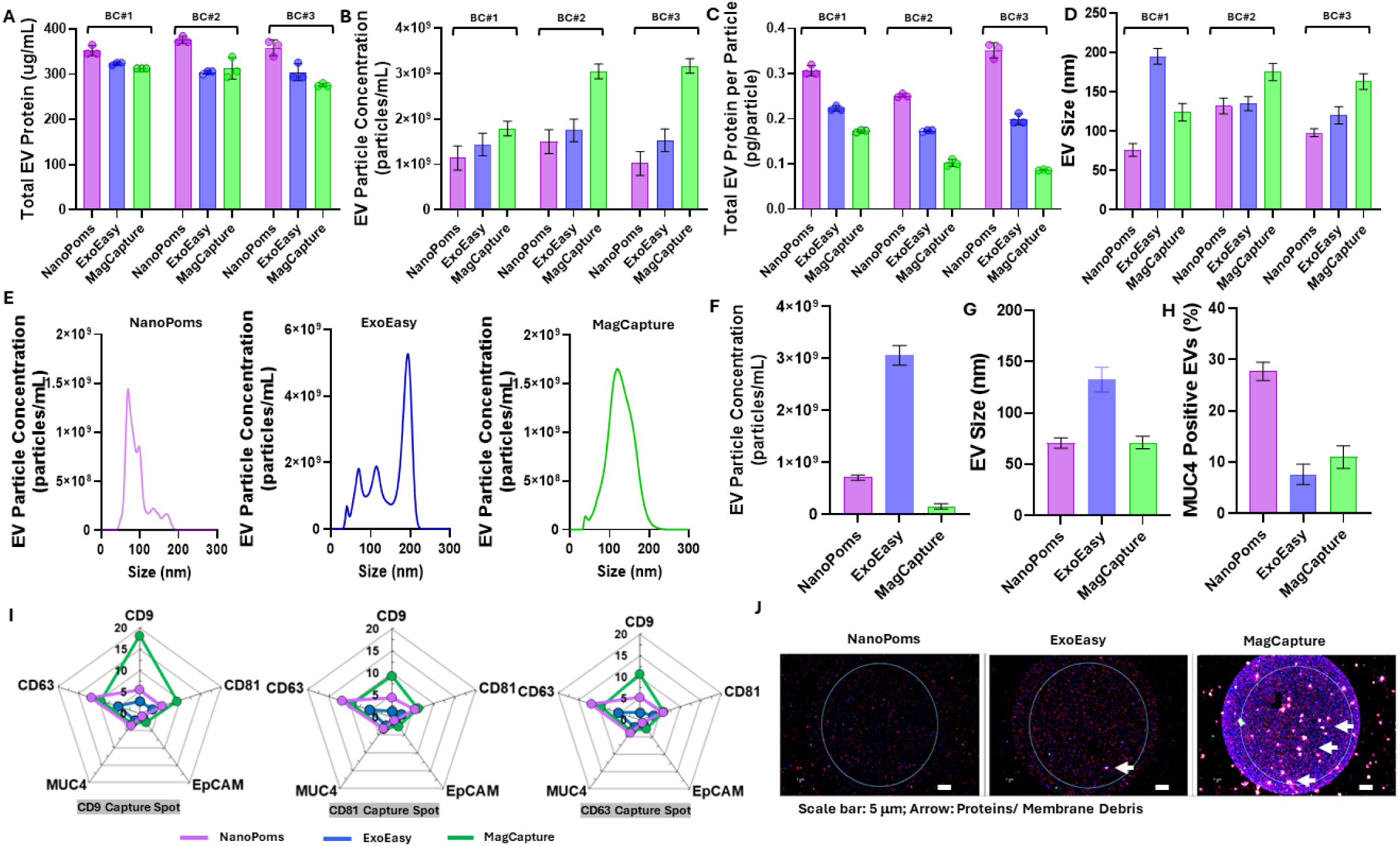
Benchmarking NanoPom’s uEV isolation. **A**. uEVs were isolated from three bladder cancer patients using different isolation methods for each same patient samples including NanoPoms^36, 37^, ExoEasy (Qiagen), and Fujifilm (MagCapture)^38^. This evaluation assessed total EV protein, as well as particle concentration (**B**), protein per particle (**C**), and EV size (**D**), respectively. **E**. Nanoparticle tracking analysis on size distribution profile associated with NanoPoms, ExoEasy, and MagCapture. **F.** Flow cytometry NanoAnalyzer (NanoFCM) characterization on EV particle concentration, size (**G**), and bladder tumor marker MUC4 expression level (**H**), using different isolation methods against the same patient sample. MUC4 is a known bladder cancer protein biomarker^39^. **I.** Spider array plots to assess the uEV’s surface protein abundance and colocalization for MUC4, EpCAM, CD81, CD9, and CD63 on the NanoView’s CD9, CD81, and CD63 capture spots. **J.** Representative images from the NanoView R100 to indicate uEV purity per isolation method. For example, MagCapture is observed to have large debris and contamination affecting the observed signal.

### uEV-based mRNA signatures for developing BCa molecular subtyping assay

To determine if NanoPom-isolated uEVs may contain mRNA markers that subtype BCa, we isolated uEVs from 16 patients to perform transcriptomic analysis and biomarker discovery (Table S1). In our discovery cohort, 6 patients were confirmed to have NMIBC and 10 with MIBC. Through differential analysis using DESeq2 (FDR < 0.05), we identified a set of mRNA biomarkers that subtyped BCa (Figure 3A), including an EV-biogenesis marker, *TSG101*, which validates that our isolated EVs are of high purity and supports the premise that EV biology is related to BCa. Next, we aimed to externally validate if our discovered EV biomarkers could subtype tumor tissue by using a ComBat-seq correction of the Xena dataset to our EV dataset. Through a supervised UMAP analysis, we verified that the individuals in the Xena dataset were strongly subtyped and concorded with patients from our discovery cohort based on their shared gene expression (Figure 3B and D). Under the premise that commercial assays including Xpert, Galeas, and CxBladder likely obtain uEV-derived mRNAs, we subtyped Xena’s dataset with their known gene sets (Figure 3C and Table S2). These known bladder cancer gene signatures showed a high tendency (>60%) for Galeas and Xpert to subtype Xena as MIBC (Figure 3E-F). On the other hand, CxBladder’s gene signature to differentiate subtypes resulted in the formation of non-homogeneous groups for MIBC and NMIBC. Unlike the homogenous groupings formed by the other gene signatures, these non-homogeneous groups identified by CxBladder are not easily separable from each other to confidently distinguish MIBC from NMIBC. We further evaluated if each mRNA signature would recapitulate their identified BCa subtypes determined by our UMAP-based analysis within the raw Xena dataset by using four machine learning models: logistic regression, linear discriminant analysis (LDA), random forest, and an ensemble model composed of these three models using a majority vote scheme. We showed that our uEV-based mRNA signature and Xpert highly recapitulated their BCa subtypes, observing standard model performance values >88%, while CxBladder and Galeas were observed at less than 80% (Table S2). However, we do observe a ∼57% concordance between our uEVs, Xpert, Galeas, and CxBladder to differentiate between MIBC and NMIBC in the Xena dataset. Overall, these results suggest that our discovered uEV biomarkers may have a strong capacity to differentiate MIBC from NMIBC.

**Figure 3.**
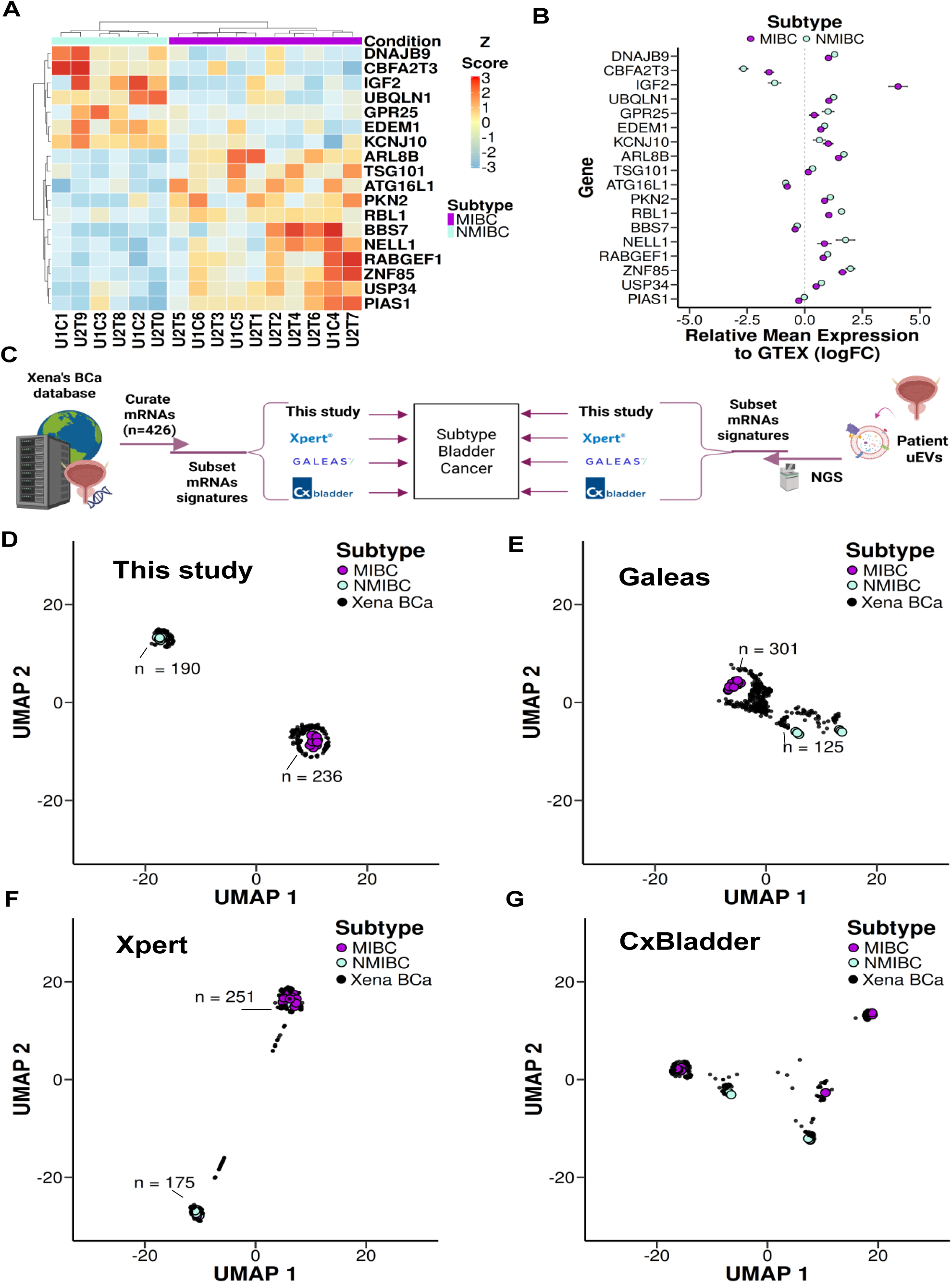
Urinary EV transcriptomic analysis to determine bladder cancer subtype biomarkers. **A**. Heatmap visualization of significant markers (FDR < 0.05) differentiating invasive and non-invasive bladder cancers. **B.** UCSC’s Xena dataset was subtyped using the discovered uEV biomarkers and nearest neighbor analysis within a uniform manifold approximation projection (UMAP) space. Relative mean expression analysis was performed by comparing the subtyped bladder cancer mRNA expression to GTEx. **C.** Visualization of how the subtyping was performed for each gene set. **D.** Discovered biomarker bladder cancer subtyping presented within the UMAP space. **E-G.** Bladder cancer subtyping analysis was performed for Galeas, Xpert, and CxBladder, respectively.

### Urinary EV biomarkers landscape for characterizing bladder cancer progression and subtypes

After determining the set of uEV-mRNAs that may distinguish MIBC from NMIBC from our transcriptomic dataset, we performed functional enrichment analysis using HumanBase’s^40^ bladder tissue network to investigate how uEVs provide information regarding gene connectivity and associated transduction specific to MIBC and NMIBC (Figure 4A). In HumanBase’s bladder tissue network, we discovered that only BBS7 was specific to MIBC, while MIBC shared RABGEF1, EDEM1, UBQLN1, and DNAJB9. On the other hand, NMIBC was specific for ZNF85, USP34, CBFA2T3, PKN2, ATG16L1, ARL8B, and IGF2. However, our analysis revealed that both NMIBC and MIBC have a direct connection to TBX3 and TP63, known as COSMIC cancer genes to undergo frequent somatic mutations. Gene ontology analysis by gProfiler (FDR < 0.05, Supplementary File 1) revealed that uEVs provided information regarding MIBC’s pathways affected by TGFB signaling, Wnt signaling, fat cell differentiation, and BBSomes. For NMIBC, we showed that more information provided by uEVs, as shown to be associated with monocyte and lymphocyte proliferation, and smooth muscle cell differentiation (Figure 4B). Both MIBC and NMIBC shared pathways associated with uEVs, further validating that our isolation method captures high-quality uEVs to differentiate NMIBC from MIBC.

**Figure 4.**
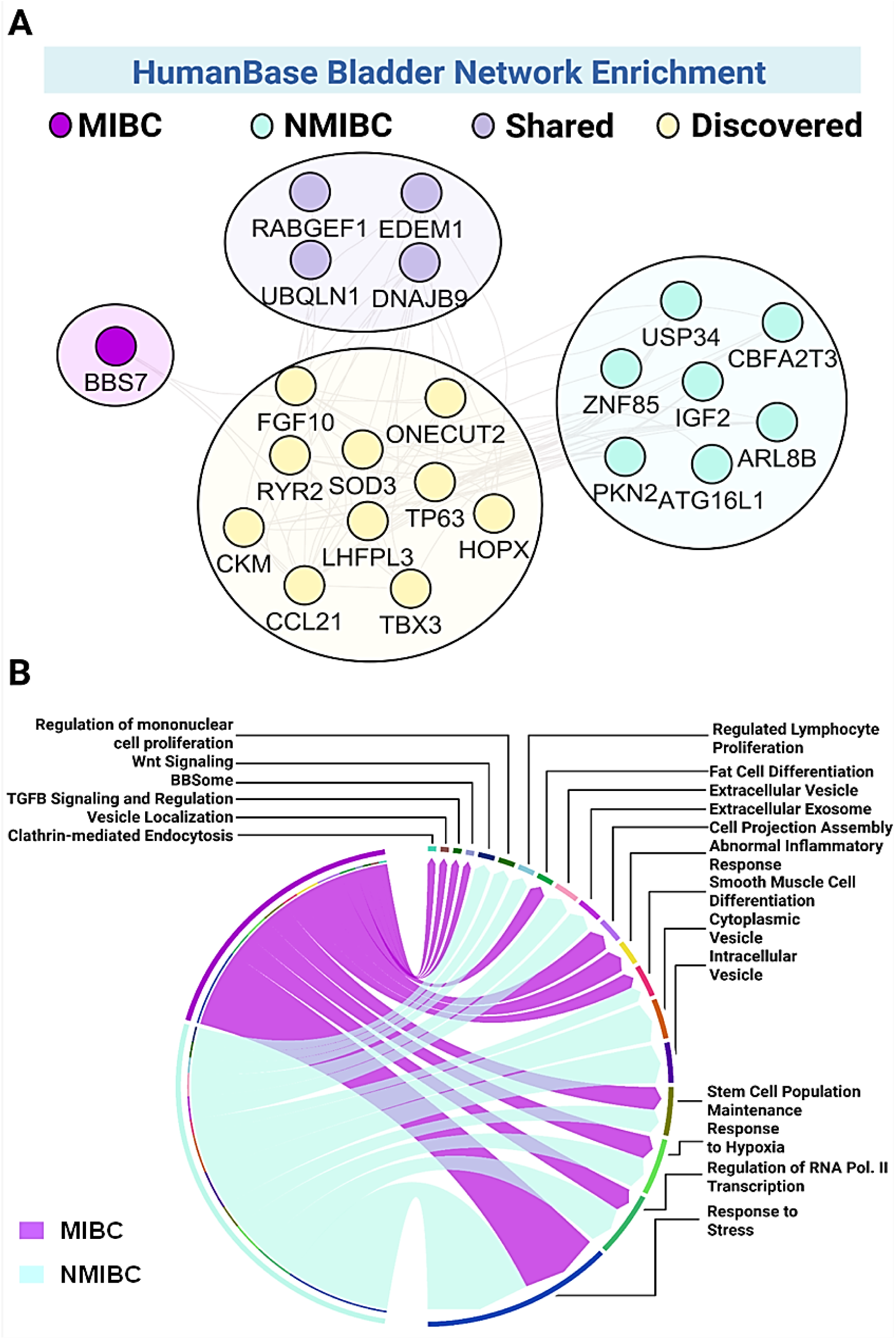
Discovered uEV mRNAs detail distinctly altered functional pathways between MIBC and NMIBC. **A**. Functional enrichment analysis was performed using HumanBase’s^28^ Bladder Tissue network to determine gene connectivity connections specific to MIBC and NMIBC. Utilizing a 0.7 confidence cutoff and removing non-connected genes, we visualized the network using Cytoscape^41^. Each node’s color corresponds to its association to MIBC, NMIBC, if the genes are shared, and discovered signaling. **B.** Functional gene ontology analysis was performed with gProfiler2^42^ (FDR < 0.05) on each gene set specific to MIBC and NMIBC, visualized as a chord plot. Each edge corresponds to the number of genes associated with the identified pathway.

### Integrating plasma and urine EV liquid biopsy for rigorous bladder cancer subtyping

Given that a patient’s circulating EVs are also transported into urine and reciprocated by subtyped bladder cancers actively secreting EVs into circulation, we hypothesized that isolating plasma EVs to evaluate their mRNA corresponding to the urinary EV signature would both improve diagnostic performance and validate our discovered biomarkers. Using the NanoPom isolation, we isolated plasma EVs from our validation cohort corresponding to MIBC, NMIBC, and healthy statuses. We found a significant difference between the strata across all EV metrics, including particle concentration (Figure 5A) and particle size (Figure 5B), total RNA (Figures 5C-D), and proteins (Figures 5E-F). However, when we integrated the plasma and urine EV transcriptomic data to observe cohort status by LDA, we observed strong clustering (Figure 5G). Given that LDA is a supervised dimensionality reduction technique that separates cohorts based on the mean and covariance of the applied gene set, rather than by statistical significance, we performed differential analysis with DESeq2 in the plasma dataset to verify LDA’s findings. Differential analysis by DESeq2 (FDR <0.05) revealed that the plasma cohort concorded well with the urine cohort, exhibiting similar mRNA signatures for DNAJB9, GPR25, CBFA2T3, IGF2, KCNJ10, ARL8B, and RABGEF, which differentiated MIBC from NMIBC (Figure 5H). Upon evaluating the biomarkers, our LDA model learned to identify each cohort. The strongest markers revealed by our biplot analysis were RBL1, NELL1, and UBQLN1 for MIBC; KCNJ10 and PIAS1 for healthy individuals; and ZNF85, GPR25, and IGF2 for NMIBC, which also agreed with our differential analysis (Figure 5I). These results suggest that our discovered biomarkers from uEVs concorded well with isolated circulating EVs, validating our uEV mRNA signature to differentiate MIBC from NMIBC.

**Figure 5.**
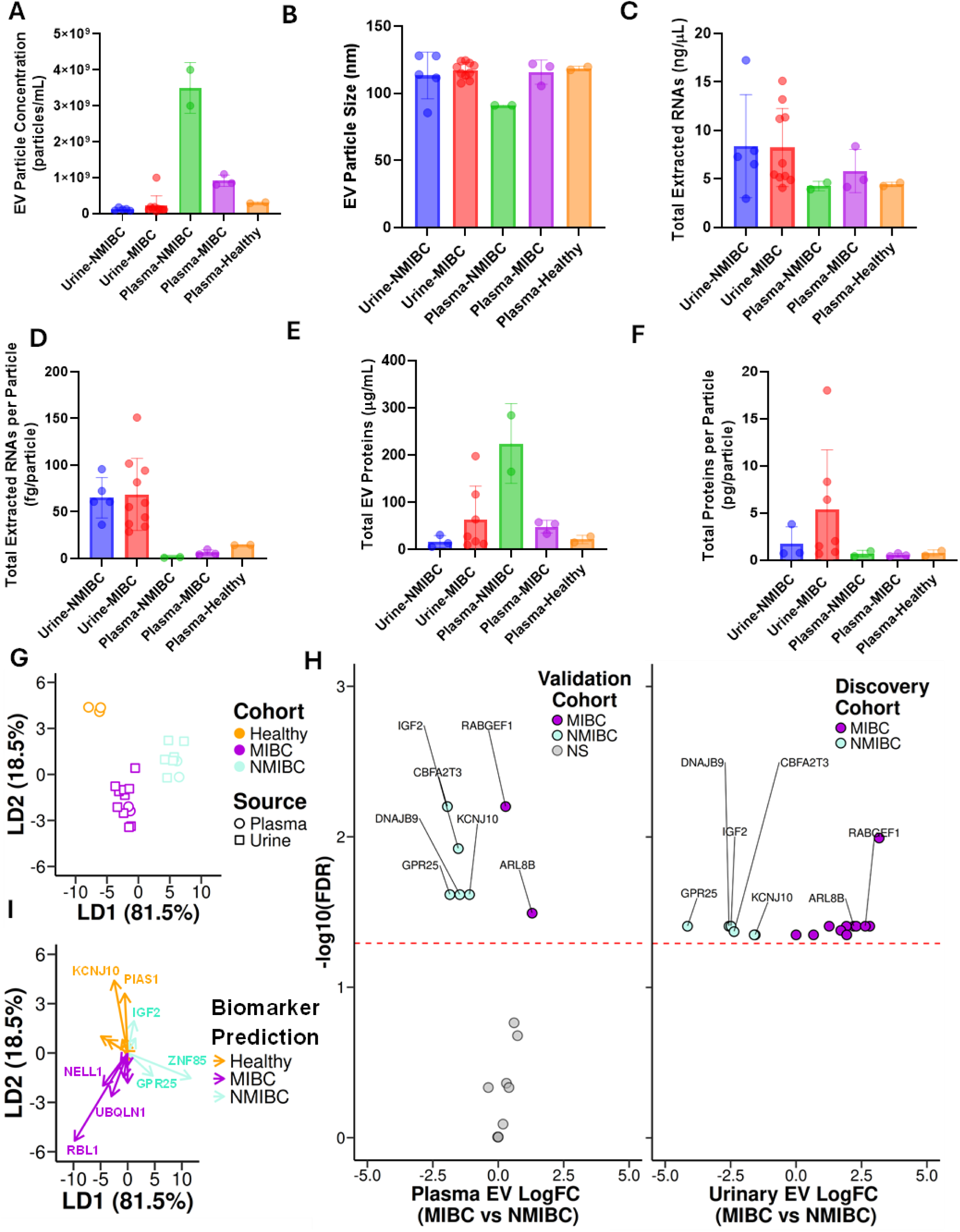
Bladder cancer subtyping validation. To validate whether the discovered bladder cancer biomarkers are present in circulating EVs, we isolated plasma EVs from different bladder cancer patients and performed transcriptomic analysis to evaluate the biomarker signature, predicting whether individuals are of bladder cancer subtypes or healthy. **A-F.** Across the strata (plasma or urine and invasive, non-invasive, or healthy), we quantified the following EV characteristics, including **A.** EV yield, **B.** EV particle size, **C.** total EV RNA, **D.** total extracted RNAs per particle, **E.** total EV proteins, and **F.** total EV protein per particle, respectively. **G**. LDA visualization of predicted cohort status using both plasma and urine EV data. **H.** Volcano plot analysis to validate the 18 biomarkers in plasma, as discovered in urine. Biomarkers were labelled if they were significant in both plasma and urine, while gray points in the validation indicate no significance. **I.** Biplot analysis of our LDA model to determine most important biomarkers learned by the model to classify healthy, MIBC, and NMIBC cohorts.

### ExoMasso for prognostic evaluation on uEV liquid biopsy for bladder cancer subtyping

Currently, uEV-based liquid biopsy has only yielded diagnostic outputs, as there do not exist any models to prognosticate patients using uEV-derived transcripts, despite databases like Xena existing with patient survival data and mRNA signatures. Thus, we built ExoMasso to address the general EV-based liquid biopsy prognostication problem using an autoencoder-based deep learning model (Figure 6A). We determined ExoMasso was optimized by evaluating when the average Wasserstein and Kolmogorov-Smirnov distances of the uEV and MIBC (Figures S1 and S2) and NMIBC mRNA signatures (Figures S3 and S4) were less than 1. The per-fold Pearson correlation coefficient (PCC) between the predicted and true mRNA signatures for MIBC and NMIBC were 0.947 (3 folds, 5 initializations, Figure S5) and 0.912 (2 folds, 5 initializations, Figure S6), indicating ExoMasso has good generalization capacity. Using the entire dataset for MIBC and NMIBC, ExoMasso generated PCC values of 0.986 and 0.985 for MIBC (Figure 6B) and NMIBC (Figure 6C), respectively. Furthermore, LDA showed that ExoMasso learns distinctive embeddings when predicting how uEV mRNAs infer MIBC and NMIBC mRNA signatures (Figure 6D). Next, we composed a Cox-Hazard model using Xena’s overall survival data and mRNA signatures, generating molecular grades corresponding to patient survival for each bladder cancer subtype identified earlier by our uEV biomarkers (Figures 6E-G). Our Cox-Hazard model enables BCa patient prognostication into three categories, low-, intermediate-, and high-grade molecular subtypes ranked by mortality. We predicted each patient’s molecular grade based on their predicted subtype, demonstrating ExoMasso’s clinical potential to identify patient risk given a bladder cancer subtyped diagnosis (Figures 6H-I).

**Figure 6.**
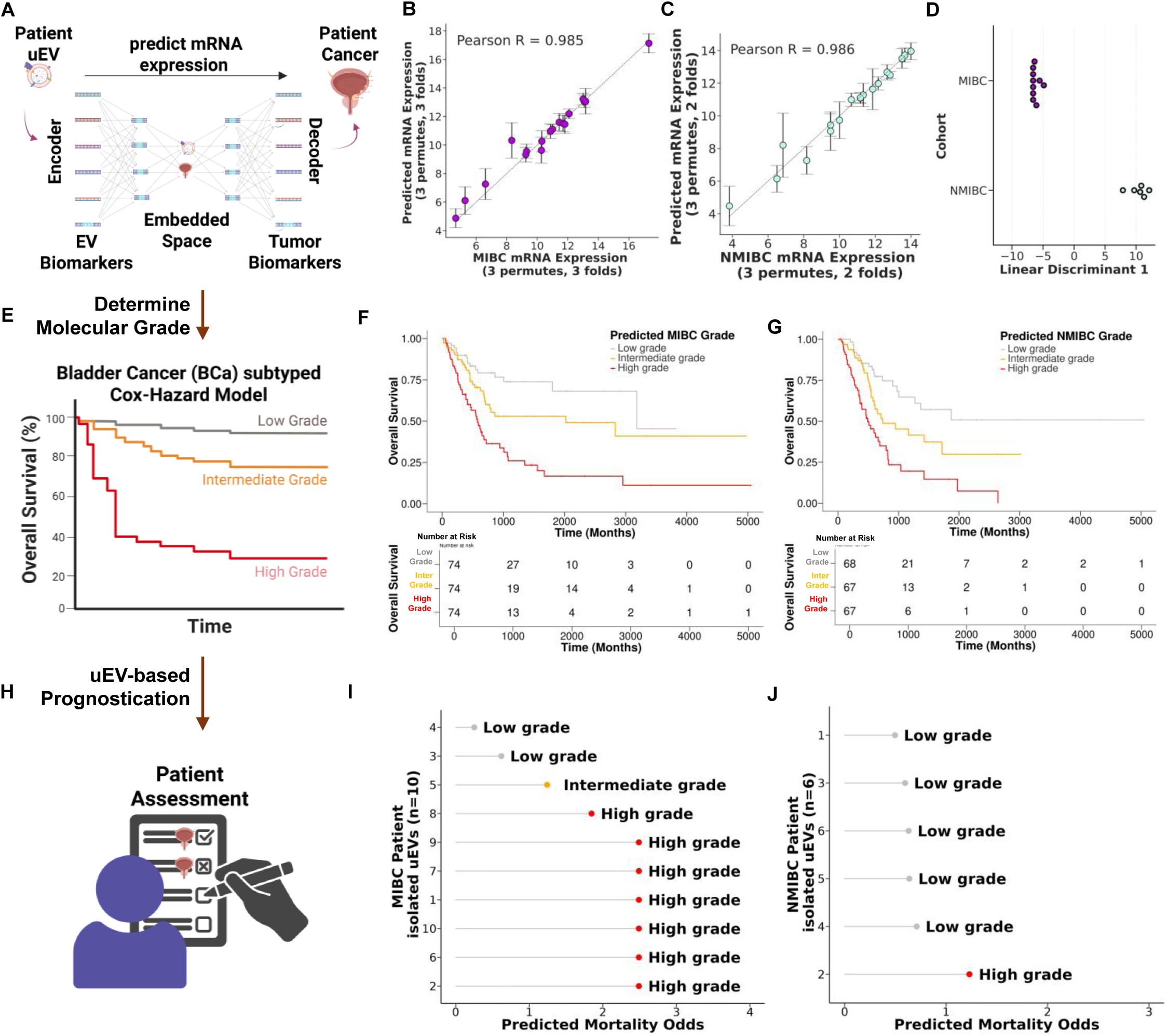
ExoMasso’s bladder cancer subtyped prognosis. **A**. Schematic detailing ExoMasso’s operation. ExoMasso is an autoencoding deep learning model built to predict mRNA expression in bladder cancers when given a set of uEV mRNAs. Upstream, our MIBC-and-NMIBC specific biomarkers are first determined, then batch corrected against the Xena dataset before introduction into ExoMasso. As patient uEVs are encoded, ExoMasso is then trained to determine the embedded space that connects the mRNA distributions between uEV and bladder cancer samples, followed by decoding out what the expected mRNA expression in bladder cancer would be given a uEV sample. **B**-**C.** Correlation plots between the predicted and true BCa expression, using grand-means across permutes and folds, after ExoMasso’s training to decode uEVs into MIBC and NMIBC, respectively. Permutes represent different seeds used to initialize ExoMasso. We used K-fold cross-validation to assess ExoMasso’s generalization, where the number of folds is listed per permute. **D.** Linear discriminant analysis was used as a dimensionality reduction technique to visualize sample similarity for uEVs predicted to be MIBC and NMIBC, respectively. **E.** A Cox-Hazard model is composed from Xena’s dataset using the uEV biomarkers, followed by quantile binning to generate molecular grades corresponding to survival. **F-G**. Cox-Hazard determined survival curves with prognostication on Xena’s dataset for MIBC and NMIBC, respectively**. H**. Patient assessment is then conducted using our Cox-Hazard model on ExoMasso predicted mRNA expression from uEVs associated to MIBC or NMIBC. **I-J.** Prognostic predictions on our patient cohort for MIBC and NMIBC respectively. The predicted mortality odds are the Cox-Hazard model’s prediction of patient mortality.

## Discussion

MIBC and NMIBC subtyping for diagnostic and prognostication is routinely done with invasive procedures including cystoscopy with biopsy for histological grading. However, cystoscopy is underpinned with significant variability in the bladder muscle examined and subsequent pathological agreeance^43^. Non-invasive procedures including CT and PET imaging^44^ and urinary cytology^45^ have high detection variation that induces significant error when determining malignant tumors upon incident findings, driving the development of urinary molecular marker tests including Galeas, CxBladder, and Xpert. Although these urinary tests are established to detect BCa, their sensitivity and specificity (<90% all) are still affected by cohort variation, necessitating new approaches to not only detect and differentiate MIBC and NMIBC, but also prognosticate BCa subtypes. Here we presented our strategy to detect, subtype, and prognosticate BCa using an AI-driven uEV-based liquid biopsy method (Figure 1).

Previous reports^46–49^ have isolated uEVs and performed biomarker discovery to differentiate NMIBC and MIBC^50^. However, these reports have not ensured that high-quality EVs were obtained to substantiate whether their discovered biomarkers can distinguish between patients with MIBC and NMIBC. Specifically, EV isolation induced heterogeneity^32, 33, 51^ is known to lead to conflicting conclusions and hinder reproducibility, impeding the development of uEV-based diagnostic and prognostic evaluations for BCa. Indeed, we showed three established isolation methods (NanoPom, ExoEasy, and FujiFilm) to obtain EVs yielded significantly different results when each isolation method was performed on the same patients (Figure 2). Indeed, we observed significant non-EV contamination with ExoEasy and Fujifilm (Figure 2j). Since urine is a highly heterogenous fluid containing thousands of expressed proteins^7^, hundreds of metabolites^8^, and tens of cell free mRNA^9^, removing non-EV contaminants by purification and utilizing an isolation method that is specific for EVs is crucial to discover unbiased biomarkers. ExoEasy (from Qiagen) utilizes membrane filtration to capture, purify, and extract EVs, while Fujifilm targets membrane phosphatidylserine using TIM4^38^. Our NanoPom-based isolation method is well-established^32, 36, 37, 52^ to capture high-quality EVs by combining the unique surface topography containing nanoscale cavities and, in this study, using photocleavable tetraspanin antibodies (CD9/CD63/CD81) for the capture-and-release of high-quality uEVs from urine and plasma.

Through next-generation sequencing of isolated uEV RNAs, we determined a uEV-based mRNA signature that distinguished MIBC from NMIBC (Figure 3A). Since uEVs are mostly derived from bladder tissue, we hypothesized that the uEV mRNA signature we discovered in our study would reflect their distinguishing capacity in next-generation sequencing bladder tissues from patients known to have BCa. We curated this data from the Xena database, which integrates all public cancer genomic data from TCGA^53^, Genomic Data Commons^54^, cBioPortal^55^, and International Cancer Genomics^56^ Consortium into tissue-specific, multi-omic cancer datasets, including transcript, lncRNA, exon, miRNA, DNA methylation, ATAC-seq, survival endpoints, and mRNA expression. However, our curated dataset lacked information on tumor stage. Additionally, there have not been any analytical methods that have attempted to evaluate the concordance in mRNA signatures between EVs and tumors. Under this premise, we addressed this analytical problem by utilizing a UMAP-based model to perform supervised dimensionality reduction and learn an embedding space that represents EV mRNA signatures, enabling the assessment of mRNA signature similarity to bladder tumors (Figure 3C). Supervised dimensionality reduction is an algorithmic technique that leverages cohort information to enhance cohort separation while finding the best principal projection axes using a weighted combination of mRNAs that best represent the data. This approach is often used in single cell sequencing to improve the differentiation between known annotated cell signatures. Our UMAP-based model would then predict the mRNA concordance between our uEV mRNAs, defining MIBC and NMIBC, and bladder cancers as a function of embedded distance. Using this approach, we demonstrated that nearly 50% of Xena samples exhibited our discovered mRNA signatures associated with MIBC and NMIBC (Figure 3D). Since urinary molecular tests likely include uEVs in their sampling, we evaluated known gene and mRNA signatures (Galeas, Xpert, and CxBladder), showing that under the uEV context, these tests have a high preference for identifying MIBC (Galeas and Xpert) or may not differentiate BCa at all (CxBladder). However, upon evaluating Xena with all mRNA signatures, we did observe a ∼57% BCa subtype concordance. Moreover, our uEV mRNA signatures are directly implicated in functional cell programming that describes NMIBC’s progression into MIBC specifically (Figure 4), identifying that alterations in smooth muscle cell differentiation, extracellular vesicle pathways, and direct links to TP63 and TBX3, genes commonly presenting somatic mutations in cancer progression^57^, further suggests uEV liquid biopsy may be used to probe differentiation mechanisms associated to NMIBCs transition into MIBC.

We further validated our uEV-based liquid biopsy by sampling circulating EVs from patients known to have MIBC and NMIBC (Figure 5). Building on previous findings and the premise that BCa-derived EVs enter circulation to affect functional pathways, we reperformed next-generation sequencing to evaluate whether these uEV-based biomarkers would have concordant mRNA signatures in plasma. Although the biological variation and technical bias between sources of EVs from plasma and urine would suggest that finding this signature is improbable, our NanoPom isolation method and transcriptomic analysis determined concordance between the signatures, enabling us to develop an LDA-based machine learning model integrating both plasma and urine to distinguish healthy individuals, and patients with MIBC, and NMIBC completely (Figure 5G). However, machine learning models do not rely on differential statistics to classify cohorts; instead, they rely on mathematical functions that distinguish means and covariances. In parallel with LDA, we demonstrated through differential analysis using DESeq2 (Figure 5H) that most of the discovered uEV biomarkers were statistically significant in plasma-derived EVs. These specific uEV biomarkers were also identified by the LDA model as the most significant for classifying MIBC, NMIBC, and healthy samples. These results validated our approach to rigorously and reproducibly determine MIBC and NMIBC in patients.

uEV-based liquid biopsy has the potential to distinguish MIBC and NMIBC and provide prognostic outcomes for patients with BCa. However, no model is currently available that can predict molecular grades and survival outcomes in BCa patients using EV-derived mRNA signatures. We developed ExoMasso to address this significant prognostic challenge (Figure 6). ExoMasso is an autoencoder deep learning model that leverages uEV mRNA data from patients classified to MIBC or NMIBC, to infer their expected BCa mRNA signature. Autoencoders utilize a mathematical framework that compresses input information into a reduced latent space, followed by uncompressing this reduced latent space into the original information. In our hands, the ExoMasso framework aims to map the mean mRNA distribution across all biomarkers between our uEV dataset and Xena’s BCa dataset by mathematically learning an embedded space shared between the two mRNA signatures (Figures 6B and 6C). ExoMasso minimizes the maximum mean discrepancy (MMD) between the ComBat-seq corrected uEV and subtyped Xena datasets, which we track at the per-gene level using the Wasserstein distance. While autoencoding frameworks do not strictly require strong prior knowledge of the joint distribution, the presence of such priors facilitates optimization by guiding the model toward a shared latent space. Furthermore, ComBat-seq helps ensure that the latent space learned by ExoMasso accurately reflects the underlying mRNA signatures that capture MIBC and NMIBC biology, rather than technical artifacts that may lead to failure in minimizing the MMD between the datasets. This latent space was initially demonstrated by our UMAP analysis, which showed that the uEV and Xena datasets occupy overlapping embedded spaces, laying the groundwork for ExoMasso to successfully minimize the MMD and predict BCa mRNA signatures using uEV data.

Since ExoMasso enabled inference into expected BCa mRNA distributions, this provided a mechanism to evaluate prognostic outcomes in sampled patients by building a Cox-Hazard regression model using the Xena transcriptomic dataset and associated survival outcomes with the subtyped labels (Figure 6E-G).We demonstrated that our uEV-based biomarkers enabled molecular grading into low, intermediate, and high-grade categories, ranked by mortality outcomes. Thus, by using the Cox-Hazard regression model, we then graded our patients who were distinctive for MIBC and NMIBC, showing that MIBC had a much higher likelihood of mortality than NMIBC, suggesting that our method for AI-driven uEV liquid biopsy may agree with clinical outcomes describing MIBC as having more aggressive and higher tumor staging than NMIBC. Our platform combines a clinically friendly EV isolation assay with transcriptomic analysis to advance towards personalized risk management and guided therapy. We expect our platform to serve as a framework to support EV-based biological conclusions and advancement towards clinically translatable outcomes.

## Data and Code Availability

All raw and processed sequencing data generated by this study is available at the NCBI Gene Expression Omnibus (GEO# GSE308996). Computer code used in this manuscript is available at github.com/zfg2013/ExoMasso. All code used in this manuscript is shared at this repository, and to ensure reproducibility, we provide a README with version information for each tool plus any parameter settings used in the data processing and analysi

## Data Availability

All raw and processed sequencing data generated by this study is available at the NCBI Gene Expression Omnibus (GEO# GSE308996). Computer code used in this manuscript is available at github.com/zfg2013/ExoMasso. All data produced in the present study are available upon reasonable request to the authors.

## Acknowledgments

The authors would like to thank the Interdisciplinary Center for Biotechnology Research (ICBR) core facility at the University of Florida for sequencing services.

## Contributions

Conceptualization: ZG, KG, PO, MH Methodology: ZG, TH, JK, KG, PO, MH Investigation: ZG, TH, JK. Visualization: ZG, PO, MH Supervision: MH Writing—original draft: ZG, PO, MH Writing—review and editing: ZG, PO, MH.

## References

1. Knollman, H.; Godwin, J. L.; Jain, R.; Wong, Y. N.; Plimack, E. R.; Geynisman, D. M., Muscle-invasive urothelial bladder cancer: an update on systemic therapy. Ther Adv Urol 2015, 7 (6), 312–30.

2. Walker, J. M.; O’Malley, P.; He, M., Applications of Exosomes in Diagnosing Muscle Invasive Bladder Cancer. Pharmaceutics 2022, 14 (10).

3. Knowles, M. A.; Hurst, C. D., Molecular biology of bladder cancer: new insights into pathogenesis and clinical diversity. Nat Rev Cancer 2015, 15 (1), 25–41.

4. Fevrier, B.; Raposo, G., Exosomes: endosomal-derived vesicles shipping extracellular messages. Curr Opin Cell Biol 2004, 16 (4), 415–21.

5. Baumgart, S.; Meschkat, P.; Edelmann, P.; Heinzelmann, J.; Pryalukhin, A.; Bohle, R.; Heinzelbecker, J.; Stockle, M.; Junker, K., MicroRNAs in tumor samples and urinary extracellular vesicles as a putative diagnostic tool for muscle-invasive bladder cancer. J Cancer Res Clin Oncol 2019, 145 (11), 2725–2736.

6. Jiang, Z.; Zhang, Y.; Zhang, Y.; Jia, Z.; Zhang, Z.; Yang, J., Cancer derived exosomes induce macrophages immunosuppressive polarization to promote bladder cancer progression. Cell Commun Signal 2021, 19 (1), 93.

7. Chang, Q.; Chen, Y.; Yin, J.; Wang, T.; Dai, Y.; Wu, Z.; Guo, Y.; Wang, L.; Zhao, Y.; Yuan, H.; Song, D.; Zhang, L., Comprehensive Urinary Proteome Profiling Analysis Identifies Diagnosis and Relapse Surveillance Biomarkers for Bladder Cancer. Journal of Proteome Research 2024, 23 (6), 2241–2252.

8. Lee, B.; Mahmud, I.; Marchica, J.; Dereziński, P.; Qi, F.; Wang, F.; Joshi, P.; Valerio, F.; Rivera, I.; Patel, V.; Pavlovich, C. P.; Garrett, T. J.; Schroth, G. P.; Sun, Y.; Perera, R. J., Integrated RNA and metabolite profiling of urine liquid biopsies for prostate cancer biomarker discovery. Scientific Reports 2020, 10 (1), 3716.

9. Kim, W. T.; Jeong, P.; Yan, C.; Kim, Y. H.; Lee, I. S.; Kang, H. W.; Kim, Y. J.; Lee, S. C.; Kim, S. J.; Kim, Y. T.; Moon, S. K.; Choi, Y. H.; Kim, I. Y.; Yun, S. J.; Kim, W. J., UBE2C cell-free RNA in urine can discriminate between bladder cancer and hematuria. Oncotarget 2016, 7 (36), 58193–58202.

10. He, N.; Thippabhotla, S.; Zhong, C.; Greenberg, Z.; Xu, L.; Pessetto, Z.; Godwin, A. K.; Zeng, Y.; He, M., Nano pom-poms prepared exosomes enable highly specific cancer biomarker detection. Commun Biol 2022, 5 (1), 660.

11. Way, G. P.; Greene, C. S., Extracting a biologically relevant latent space from cancer transcriptomes with variational autoencoders. *Pacific Symposium on Biocomputing*. Pacific Symposium on Biocomputing 2018, 23, 80–91.

12. Gretton, A.; Borgwardt, K. M.; Rasch, M. J.; Schölkopf, B.; Smola, A., A kernel two-sample test. J. Mach. Learn. Res. 2012, 13 (null), 723–773.

13. Goldman, M. J.; Craft, B.; Hastie, M.; Repečka, K.; McDade, F.; Kamath, A.; Banerjee, A.; Luo, Y.; Rogers, D.; Brooks, A. N.; Zhu, J.; Haussler, D., Visualizing and interpreting cancer genomics data via the Xena platform. Nature Biotechnology 2020, 38 (6), 675–678.

14. Cox, D. R., Regression Models and Life-Tables. Journal of the Royal Statistical Society: Series B (Methodological) 1972, 34 (2), 187–202.

15. Gordon, N. S.; McGuigan, E. K.; Ondasova, M.; Knight, J.; Baxter, L. A.; Ott, S.; Hastings, R. K.; Zeegers, M. P.; James, N. D.; Cheng, K. K.; Goel, A.; Yu, M.; Arnold, R.; Bryan, R. T.; Ward, D. G., Comparison and combination of mutation and methylation-based urine tests for bladder cancer detection. (2050-7771 (Print)).

16. Ward, D. G.; Baxter, L.; Ott, S.; Gordon, N. S.; Wang, J.; Patel, P.; Piechocki, K.; Silcock, L.; Sale, C.; Zeegers, M. P.; Cheng, K. K.; James, N. D.; Bryan, R. T., Highly Sensitive and Specific Detection of Bladder Cancer via Targeted Ultra-deep Sequencing of Urinary DNA. Eur Urol Oncol 2023, 6 (1), 67–75.

17. Li, K. D.; Chu, C. E.; Patel, M.; Meng, M. V.; Morgan, T. M.; Porten, S. P., Cxbladder Monitor testing to reduce cystoscopy frequency in patients with bladder cancer. Urologic Oncology: Seminars and Original Investigations 2023, 41 (7), 326.e1-326.e8.

18. Lotan, Y.; O’Sullivan, P.; Raman, J. D.; Shariat, S. F.; Kavalieris, L.; Frampton, C.; Guilford, P.; Luxmanan, C.; Suttie, J.; Crist, H.; Scherr, D.; Asroff, S.; Goldfischer, E.; Thill, J.; Darling, D., Clinical comparison of noninvasive urine tests for ruling out recurrent urothelial carcinoma. Urologic Oncology: Seminars and Original Investigations 2017, 35 (8), 531.e15-531.e22.

19. Sordelli, F.; Desai, A.; Dagnino, F.; Contieri, R.; Giuriolo, S.; Paciotti, M.; Fasulo, V.; Mancon, S.; Maffei, D.; Avolio, P. P.; Da Rin, G.; Maura, F.; Vanni, E.; Federico, D.; Colombo, P.; Lughezzani, G.; Buffi, N. M.; Casale, P.; Saita, A.; Lazzeri, M.; Hurle, R.; Voza, A., Xpert Bladder Cancer Detection in Emergency Setting Assessment (XESA Project): A Prospective, Single-centre Trial. Eur Urol Open Sci 2025, 71, 172–179.

20. Sharma, G.; Sharma, A.; Krishna, M.; Devana, S. K.; Singh, S. K., Xpert bladder cancer monitor in surveillance of bladder cancer: Systematic review and meta-analysis. Urol Oncol 2022, 40 (4), 163.e1-163.e9.

21. Graim, K.; Gorenshteyn, D.; Robinson, D. G.; Carriero, N. J.; Cahill, J. A.; Chakrabarti, R.; Goldschmidt, M. H.; Durham, A. C.; Funk, J.; Storey, J. D.; Kristensen, V. N.; Theesfeld, C. L.; Sorenmo, K. U.; Troyanskaya, O. G., Modeling molecular development of breast cancer in canine mammary tumors. Genome Res 2020, 31 (2), 337–47.

22. Chen, S.; Zhou, Y.; Chen, Y.; Gu, J., fastp: an ultra-fast all-in-one FASTQ preprocessor. Bioinformatics 2018, 34 (17), i884–i890.

23. Patro, R.; Duggal, G.; Love, M. I.; Irizarry, R. A.; Kingsford, C., Salmon provides fast and bias-aware quantification of transcript expression. Nature Methods 2017, 14 (4), 417–419.

24. Soneson, C.; Love, M.; Robinson, M., Differential analyses for RNA-seq: transcript-level estimates improve gene-level inferences [version 1; peer review: 2 approved]. F1000Research 2015, 4 (1521).

25. Zhang, Y.; Parmigiani, G.; Johnson, W. E., ComBat-seq: batch effect adjustment for RNA-seq count data. NAR genomics and bioinformatics 2020, 2 (3), lqaa078.

26. Love, M. I.; Huber, W.; Anders, S., Moderated estimation of fold change and dispersion for RNA-seq data with DESeq2. Genome Biology 2014, 15 (12), 550.

27. Kolberg, L.; Raudvere, U.; Kuzmin, I.; Adler, P.; Vilo, J.; Peterson, H., g:Profiler—interoperable web service for functional enrichment analysis and gene identifier mapping (2023 update). Nucleic Acids Research 2023, 51 (W1), W207–W212.

28. Krishnan, A.; Zhang, R.; Yao, V.; Theesfeld, C. L.; Wong, A. K.; Tadych, A.; Volfovsky, N.; Packer, A.; Lash, A.; Troyanskaya, O. G., Genome-wide prediction and functional characterization of the genetic basis of autism spectrum disorder. Nature neuroscience 2016, 19 (11), 1454–1462.

29. Fukushima, K., Visual Feature Extraction by a Multilayered Network of Analog Threshold Elements. IEEE Transactions on Systems Science and Cybernetics 1969, 5 (4), 322–333.

30. Kingma, D. P.; Ba, J., Adam: A Method for Stochastic Optimization. CoRR 2014, *abs/1412*.6980.

31. Benjamini, Y.; Hochberg, Y., Controlling the False Discovery Rate: A Practical and Powerful Approach to Multiple Testing. Journal of the Royal Statistical Society. Series B (Methodological) 1995, 57 (1), 289–300.

32. Greenberg, Z. F.; Ali, S.; Brock, A.; Jiang, J.; Schmittgen, T. D.; Han, S.; Hughes, S. J.; Graim, K. S.; He, M., Nanomaterial isolated extracellular vesicles enable high precision identification of tumor biomarkers for pancreatic cancer liquid biopsy. Journal of Nanobiotechnology 2025, 23 (1), 467.

33. Veerman, R. E.; Teeuwen, L.; Czarnewski, P.; Güclüler Akpinar, G.; Sandberg, A.; Cao, X.; Pernemalm, M.; Orre, L. M.; Gabrielsson, S.; Eldh, M., Molecular evaluation of five different isolation methods for extracellular vesicles reveals different clinical applicability and subcellular origin. J Extracell Vesicles 2021, 10 (9), e12128.

34. Kowal, J.; Arras, G.; Colombo, M.; Jouve, M.; Morath, J. P.; Primdal-Bengtson, B.; Dingli, F.; Loew, D.; Tkach, M.; Théry, C., Proteomic comparison defines novel markers to characterize heterogeneous populations of extracellular vesicle subtypes. Proc Natl Acad Sci U S A 2016, 113 (8), E968–77.

35. Breitwieser, K.; Koch, L. F.; Tertel, T.; Proestler, E.; Burgers, L. D.; Lipps, C.; Adjaye, J.; Fürst, R.; Giebel, B.; Saul, M. J., Detailed Characterization of Small Extracellular Vesicles from Different Cell Types Based on Tetraspanin Composition by ExoView R100 Platform. Int J Mol Sci 2022, 23 (15).

36. Greenberg, Z. F.; Oshins, R.; Serban, K.; Bazargani, S. F.; Garrett, T. J.; Casanova, N. G.; Garcia, J. G. N.; He, M.; Khodayari, N., Neutrophil-derived extracellular vesicles in the plasma of alpha-1 antitrypsin deficient individuals reveal pro-inflammatory metabolic and transcriptomic signatures. Extracellular Vesicle 2025, 6, 100093.

37. Nan He, S. T., Cuncong Zhong, Zachary Greenberg, Liang Xu, Ziyan Pessetto, Andrew K. Godwin, Yong Zeng, Mei He, Nano Pom-poms Prepared Exosomes enable Highly Specific Cancer Biomarker Detection. bioRxiv 2021.

38. Yoshida, T.; Ishidome, T.; Hanayama, R., High Purity Isolation and Sensitive Quantification of Extracellular Vesicles Using Affinity to TIM4. Current Protocols in Cell Biology 2017, 77 (1), 3.45.1-3.45.18.

39. Yang, H.; Song, H.; Yip, E.; Gilpatrick, T.; Chang, K.; Allegakoen, P.; Lu, K. L.; Hui, K.; Pham, J. H.; Kasap, C.; Kumar, V.; Gayle, J.; Stohr, B. A.; Cornelia Ding, C. K.; Wiita, A. P.; Meng, M. V.; Chou, J.; Porten, S. P.; Huang, F. W., Bladder cancer variants share aggressive features including a CA125+ cell state and targetable TM4SF1 expression. Nat Commun 2025, 16 (1), 5312.

40. Greene, C. S.; Krishnan, A.; Wong, A. K.; Ricciotti, E.; Zelaya, R. A.; Himmelstein, D. S.; Zhang, R.; Hartmann, B. M.; Zaslavsky, E.; Sealfon, S. C.; Chasman, D. I.; FitzGerald, G. A.; Dolinski, K.; Grosser, T.; Troyanskaya, O. G., Understanding multicellular function and disease with human tissue-specific networks. Nature Genetics 2015, 47 (6), 569–576.

41. Shannon, P.; Markiel, A.; Ozier, O.; Baliga, N. S.; Wang, J. T.; Ramage, D.; Amin, N.; Schwikowski, B.; Ideker, T., Cytoscape: a software environment for integrated models of biomolecular interaction networks. Genome Res 2003, 13 (11), 2498–504.

42. Kolberg, L.; Raudvere, U.; Kuzmin, I.; Vilo, J.; Peterson, H., gprofiler2 -- an R package for gene list functional enrichment analysis and namespace conversion toolset g:Profiler. F1000Research 2020, 9.

43. van Straten, C.; Bruins, M. H.; Dijkstra, S.; Cornel, E. B.; Kortleve, M. D. H.; de Vocht, T. F.; Kiemeney, L.; van der Heijden, A. G., The accuracy of cystoscopy in predicting muscle invasion in newly diagnosed bladder cancer patients. World J Urol 2023, 41 (7), 1829–1835.

44. Malayeri, A. A.; Pattanayak, P.; Apolo, A. B., Imaging muscle-invasive and metastatic urothelial carcinoma. Curr Opin Urol 2015, 25 (5), 441–8.

45. Pan, T.; Lehman, E.; Raman, J. D., Performance characteristics of urinary cytology in patients presenting with gross and microscopic hematuria. Am J Clin Exp Urol 2021, 9 (5), 384–389.

46. Murakami, T.; Yamamoto, C. M.; Akino, T.; Tanaka, H.; Fukuzawa, N.; Suzuki, H.; Osawa, T.; Tsuji, T.; Seki, T.; Harada, H., Bladder cancer detection by urinary extracellular vesicle mRNA analysis. Oncotarget 2018, 9 (67), 32810–32821.

47. Sun, N.; Zhang, Z.; Yang, X.; Li, J.; Li, Q.; Kang, J.; Wei, Y.; Yu, X.; Du, R.; Hong, X.; Liu, G.; Gao, H.; Liu, D., Unveiling urinary extracellular vesicle mRNA signature for early diagnosis and prognosis of bladder cancer. Theranostics 2025, 15 (4), 1272–1284.

48. Hinestrosa, J. P.; Kurzrock, R.; Lewis, J. M.; Schork, N. J.; Schroeder, G.; Kamat, A. M.; Lowy, A. M.; Eskander, R. N.; Perrera, O.; Searson, D.; Rastegar, K.; Hughes, J. R.; Ortiz, V.; Clark, I.; Balcer, H. I.; Arakelyan, L.; Turner, R.; Billings, P. R.; Adler, M. J.; Lippman, S. M.; Krishnan, R., Early-stage multi-cancer detection using an extracellular vesicle protein-based blood test. Commun Med (Lond) 2022, 2, 29.

49. Dong, L.; Feng, M.; Kuczler, M. D.; Horie, K.; Kim, C. J.; Ma, Z.; Lombardo, K.; Lyons, H.; Amend, S. R.; Kates, M.; Bivalacqua, T. J.; McConkey, D.; Xue, W.; Choi, W.; Pienta, K. J., Tumour tissue-derived small extracellular vesicles reflect molecular subtypes of bladder cancer. J Extracell Vesicles 2024, 13 (2), e12402.

50. Huang, K.; Yang, C.; Xu, Y.; Wang, Y., Unraveling the multifaceted roles of extracellular vesicles in bladder cancer: diagnostic insights and therapeutic opportunities. Front Oncol 2025, 15, 1554819.

51. Kowal, J.; Arras, G.; Colombo, M.; Jouve, M.; Morath, J. P.; Primdal-Bengtson, B.; Dingli, F.; Loew, D.; Tkach, M.; Théry, C., Proteomic comparison defines novel markers to characterize heterogeneo us populations of extracellular vesicle subtypes. Proceedings of the National Academy of Sciences 113, E968–E977.

52. Hong, S.; Ruan, S.; Greenberg, Z.; He, M.; McGill, J. L., Development of surface engineered antigenic exosomes as vaccines for respiratory syncytial virus. Sci Rep 2021, 11 (1), 21358.

53. Weinstein, J. N.; Collisson, E. A.; Mills, G. B.; Shaw, K. R.; Ozenberger, B. A.; Ellrott, K.; Shmulevich, I.; Sander, C.; Stuart, J. M., The Cancer Genome Atlas Pan-Cancer analysis project. Nat Genet 2013, 45 (10), 1113–20.

54. Grossman, R. L.; Heath, A. P.; Ferretti, V.; Varmus, H. E.; Lowy, D. R.; Kibbe, W. A.; Staudt, L. M., Toward a Shared Vision for Cancer Genomic Data. N Engl J Med 2016, 375 (12), 1109–12.

55. Cerami, E.; Gao, J.; Dogrusoz, U.; Gross, B. E.; Sumer, S. O.; Aksoy, B. A.; Jacobsen, A.; Byrne, C. J.; Heuer, M. L.; Larsson, E.; Antipin, Y.; Reva, B.; Goldberg, A. P.; Sander, C.; Schultz, N., The cBio cancer genomics portal: an open platform for exploring multidimensional cancer genomics data. Cancer Discov 2012, 2 (5), 401–4.

56. Zhang, J.; Bajari, R.; Andric, D.; Gerthoffert, F.; Lepsa, A.; Nahal-Bose, H.; Stein, L. D.; Ferretti, V., The International Cancer Genome Consortium Data Portal. Nat Biotechnol 2019, 37 (4), 367–369.

57. Tate, J. G.; Bamford, S.; Jubb, H. C.; Sondka, Z.; Beare, D. M.; Bindal, N.; Boutselakis, H.; Cole, C. G.; Creatore, C.; Dawson, E.; Fish, P.; Harsha, B.; Hathaway, C.; Jupe, S. C.; Kok, C. Y.; Noble, K.; Ponting, L.; Ramshaw, C. C.; Rye, C. E.; Speedy, H. E.; Stefancsik, R.; Thompson, S. L.; Wang, S.; Ward, S.; Campbell, P. J.; Forbes, S. A., COSMIC: the Catalogue Of Somatic Mutations In Cancer. Nucleic Acids Res 2019, 47 (D1), D941–d947.

