## Supplementary Materials for "Urinary extracellular vesicles for high-precision bladder cancer subtyping and prognosis"

| SpecimenType | Patient_ID | Condition | Batch | Location |
|---|---|---|---|---|
| Urine | A00L | NMIBC | 1 | Clinic |
| Urine | A001 | NMIBC | 1 | Clinic |
| Urine | A004 | NMIBC | 1 | Clinic |
| Urine | B001 | MIBC | 1 | Clinic |
| Urine | B002 | MIBC | 1 | Clinic |
| Urine | B003 | MIBC | 1 | Clinic |
| Urine | A008 | MIBC | 2 | Clinic |
| Urine | A0015 | MIBC | 2 | Clinic |
| Urine | A0016 | MIBC | 2 | Clinic |
| Urine | B004 | MIBC | 2 | Clinic |
| Urine | B005 | MIBC | 2 | Clinic |
| Urine | B007 | MIBC | 2 | Clinic |
| Urine | B009 | MIBC | 2 | Clinic |
| Urine | A005 | NMIBC | 2 | Clinic |
| Urine | A006 | NMIBC | 2 | Clinic |
| Urine | A007 | NMIBC | 2 | Clinic |

**Table S1.** Summary table of patient demographics

**Logistic Regression with L2 Penalty (6 Folds)**

| | Balanced Accuracy | MCC | Precision | F1 | Sensitivity | Specificity | AUROC |
|---|---|---|---|---|---|---|---|
| **This Study** | 0.934 ± 0.014 | 0.868 ± 0.029 | 0.923 ± 0.034 | 0.927 ± 0.016 | 0.932 ± 0.024 | 0.937 ± 0.031 | 0.986 ± 0.009 |
| **Xpert** | 0.963 ± 0.031 | 0.928 ± 0.066 | 0.961 ± 0.053 | 0.957 ± 0.038 | 0.954 ± 0.036 | 0.972 ± 0.038 | 0.995 ± 0.006 |
| **Galeas** | 0.720 ± 0.040 | 0.407 ± 0.070 | 0.513 ± 0.040 | 0.600 ± 0.047 | 0.729 ± 0.087 | 0.711 ± 0.048 | 0.829 ± 0.036 |
| **Cxbladder** | 0.806 ± 0.061 | 0.580 ± 0.120 | 0.641 ± 0.084 | 0.715 ± 0.078 | 0.814 ± 0.092 | 0.798 ± 0.060 | 0.843 ± 0.058 |

**Linear Discriminant Analysis (LDA) (6 Folds)**

| | Balanced Accuracy | MCC | Precision | F1 | Sensitivity | Specificity | AUROC |
|---|---|---|---|---|---|---|---|
| **This Study** | 0.920 ± 0.020 | 0.838 ± 0.044 | 0.884 ± 0.048 | 0.911 ± 0.024 | 0.942 ± 0.023 | 0.899 ± 0.045 | 0.979 ± 0.010 |
| **Xpert** | 0.937 ± 0.032 | 0.890 ± 0.059 | 0.981 ± 0.032 | 0.930 ± 0.038 | 0.886 ± 0.052 | 0.988 ± 0.020 | 0.994 ± 0.007 |
| **Galeas** | 0.671 ± 0.030 | 0.377 ± 0.065 | 0.619 ± 0.073 | 0.526 ± 0.051 | 0.464 ± 0.067 | 0.877 ± 0.046 | 0.831 ± 0.025 |
| **Cxbladder** | 0.719 ± 0.081 | 0.497 ± 0.170 | 0.752 ± 0.141 | 0.605 ± 0.133 | 0.512 ± 0.133 | 0.926 ± 0.046 | 0.841 ± 0.056 |

**Random Forest (6 Folds)**

| | Balanced Accuracy | MCC | Precision | F1 | Sensitivity | Specificity | AUROC |
|---|---|---|---|---|---|---|---|
| **This Study** | 0.903 ± 0.037 | 0.812 ± 0.065 | 0.914 ± 0.031 | 0.891 ± 0.043 | 0.873 ± 0.079 | 0.932 ± 0.031 | 0.966 ± 0.029 |
| **Xpert** | 0.944 ± 0.056 | 0.899 ± 0.104 | 0.974 ± 0.048 | 0.936 ± 0.066 | 0.903 ± 0.090 | 0.984 ± 0.029 | 0.989 ± 0.012 |
| **Galeas** | 0.634 ± 0.037 | 0.347 ± 0.073 | 0.686 ± 0.108 | 0.441 ± 0.081 | 0.337 ± 0.095 | 0.930 ± 0.041 | 0.827 ± 0.052 |
| **Cxbladder** | 0.826 ± 0.073 | 0.679 ± 0.135 | 0.822 ± 0.089 | 0.765 ± 0.103 | 0.720 ± 0.128 | 0.933 ± 0.033 | 0.924 ± 0.026 |

**Ensemble Voting (All Models) (6 Folds)**

| | Balanced Accuracy | MCC | Precision | F1 | Sensitivity | Specificity | AUROC |
|---|---|---|---|---|---|---|---|
| **This Study** | 0.930 ± 0.015 | 0.859 ± 0.031 | 0.913 ± 0.033 | 0.922 ± 0.017 | 0.932 ± 0.024 | 0.928 ± 0.029 | 0.985 ± 0.008 |
| **Xpert** | 0.961 ± 0.036 | 0.928 ± 0.071 | 0.976 ± 0.043 | 0.956 ± 0.043 | 0.937 ± 0.050 | 0.984 ± 0.029 | 0.995 ± 0.007 |
| **Galeas** | 0.688 ± 0.042 | 0.400 ± 0.082 | 0.615 ± 0.075 | 0.554 ± 0.073 | 0.512 ± 0.095 | 0.864 ± 0.049 | 0.841 ± 0.032 |
| **Cxbladder** | 0.817 ± 0.079 | 0.649 ± 0.154 | 0.779 ± 0.122 | 0.747 ± 0.113 | 0.728 ± 0.140 | 0.905 ± 0.061 | 0.890 ± 0.044 |

**Table S2**. Summary table of machine learning algorithms used to predict MIBC and NMIBC after our UMAP-based generation of subtype labels.

| UMAP (UWOT) Parameters | Values |
| --- | --- |
| n_neighbors | 5 |
| n_epochs | 1000 |
| min_dist | 0.05 |
| metric | correlation |
| init | spectral |
| nn_method | nndescent |
| seed | 2385325 |
| target | BCa Subtype |
| dens_scale | 0.2 |

**Table S3**. The list of UMAP parameters used in this manuscript.

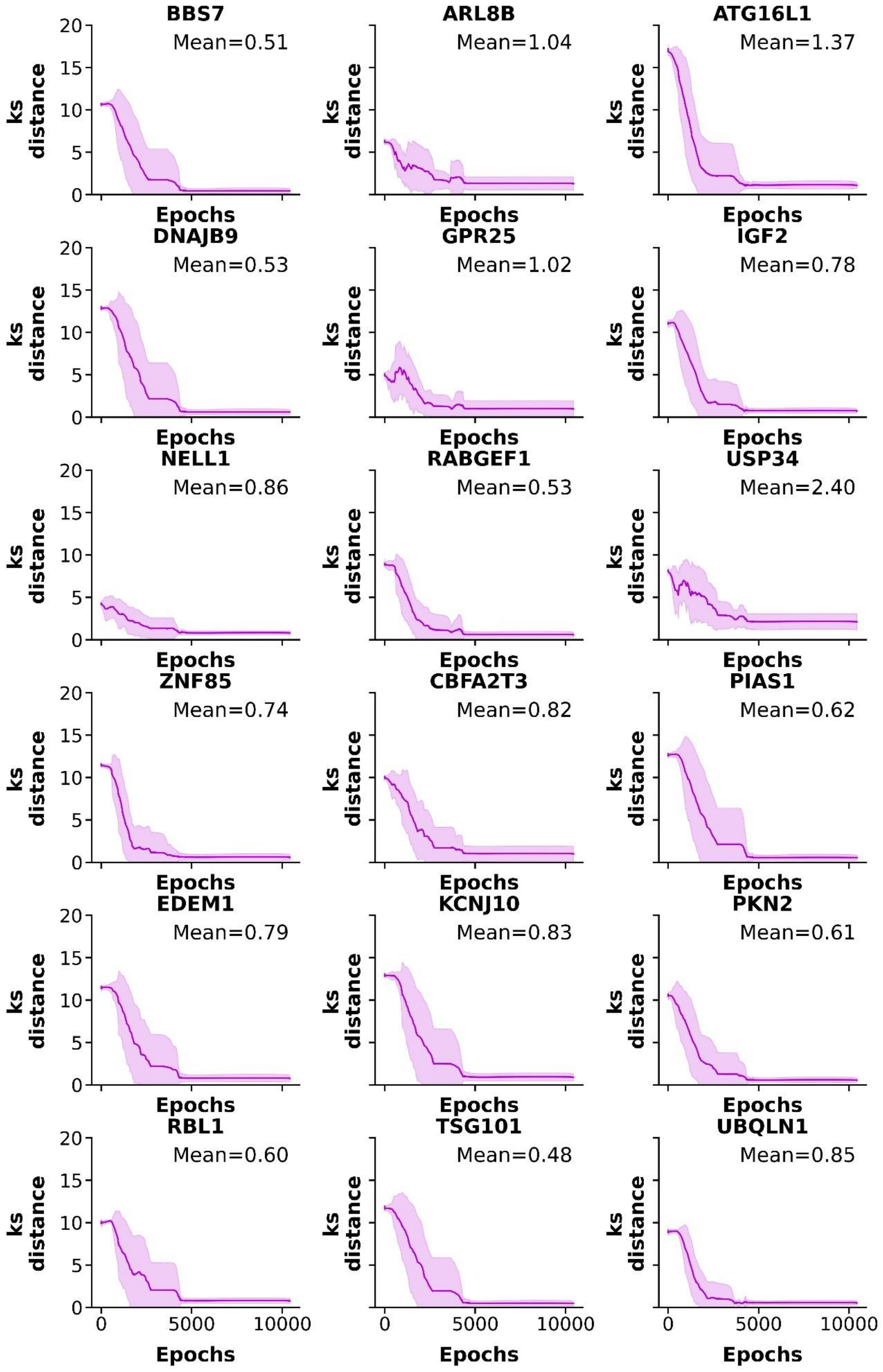


**Figure S1. ExoMasso's training optimization by of the per-mRNA Kolmogorov-Smirnov (KS) distance.** We computed the per-gene KS distance per fold per initialized model (3 models, 3 folds) between MIBC and uEV during ExoMasso's K-fold cross validation to evaluate if ExoMasso has completed training to map uEV mRNA signatures to MIBC mRNA signatures.

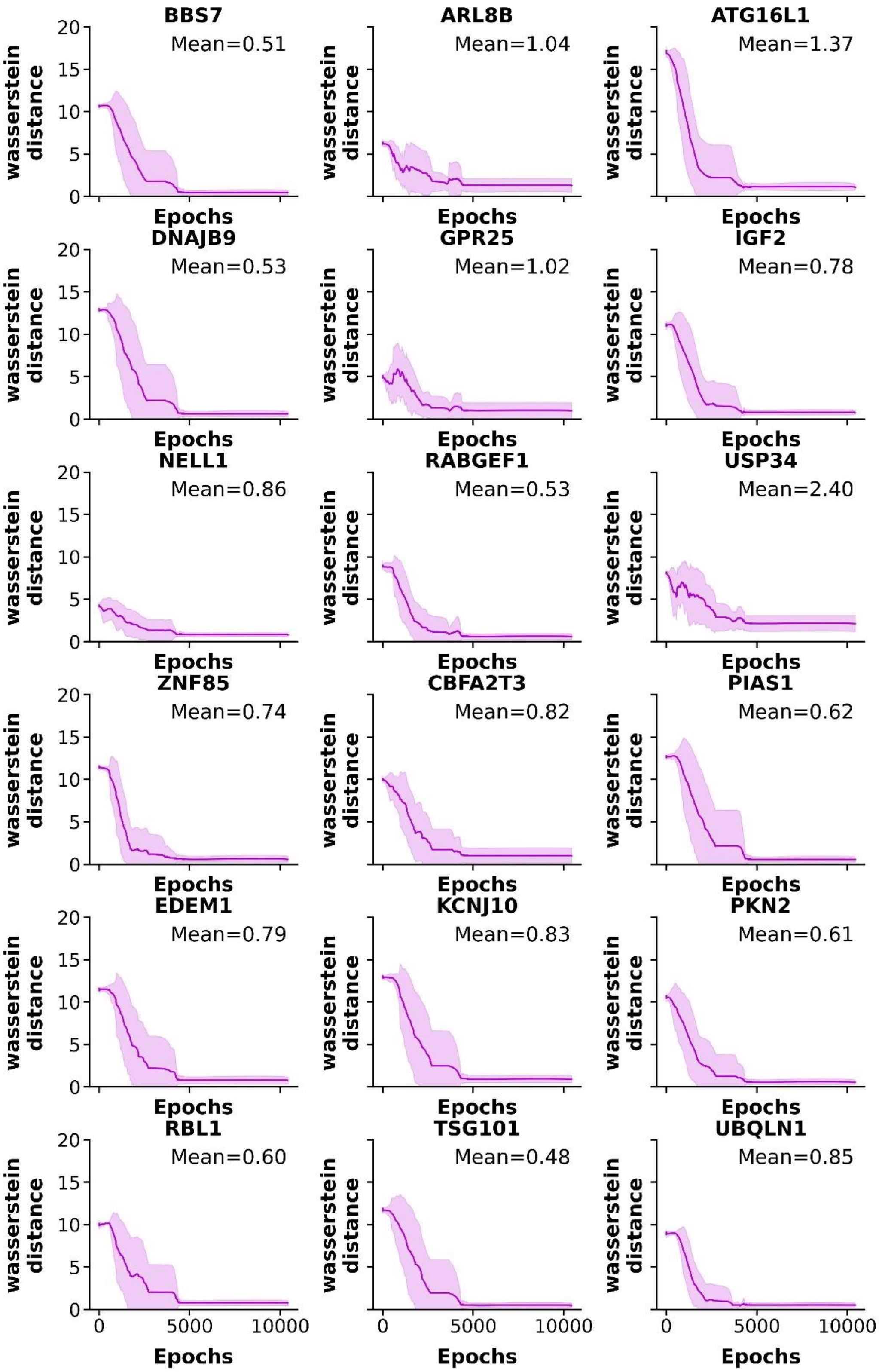


**Figure S2. ExoMasso's training optimization by of the per-mRNA Wasserstein distance.** We computed the per-gene Wasserstein distance per fold per initialized model (3 models, 3 folds) between Xena and uEV during ExoMasso's K-fold cross validation to evaluate if ExoMasso has completed training to map uEV mRNA signatures to MIBC mRNA signatures.

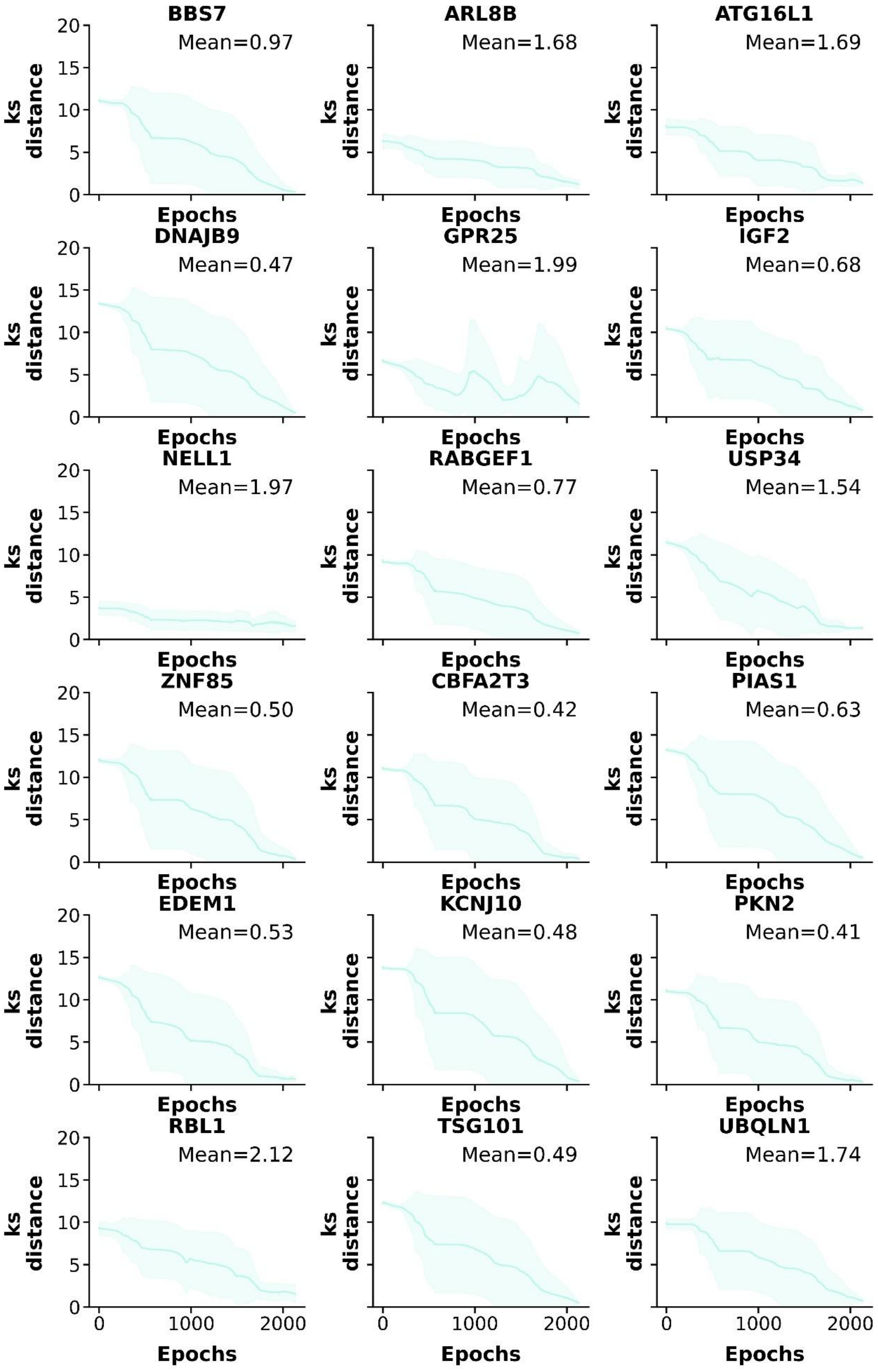


**Figure S3. ExoMasso's training optimization by the per-mRNA Kolmogorov-Smirnov (KS) distance.** We computed the per-gene KS distance per fold per initialized model (3 models, 2 folds) between NMIBC and uEV during ExoMasso's K-fold cross validation to evaluate if ExoMasso has completed training to map uEV mRNA signatures to NMIBC mRNA signatures.

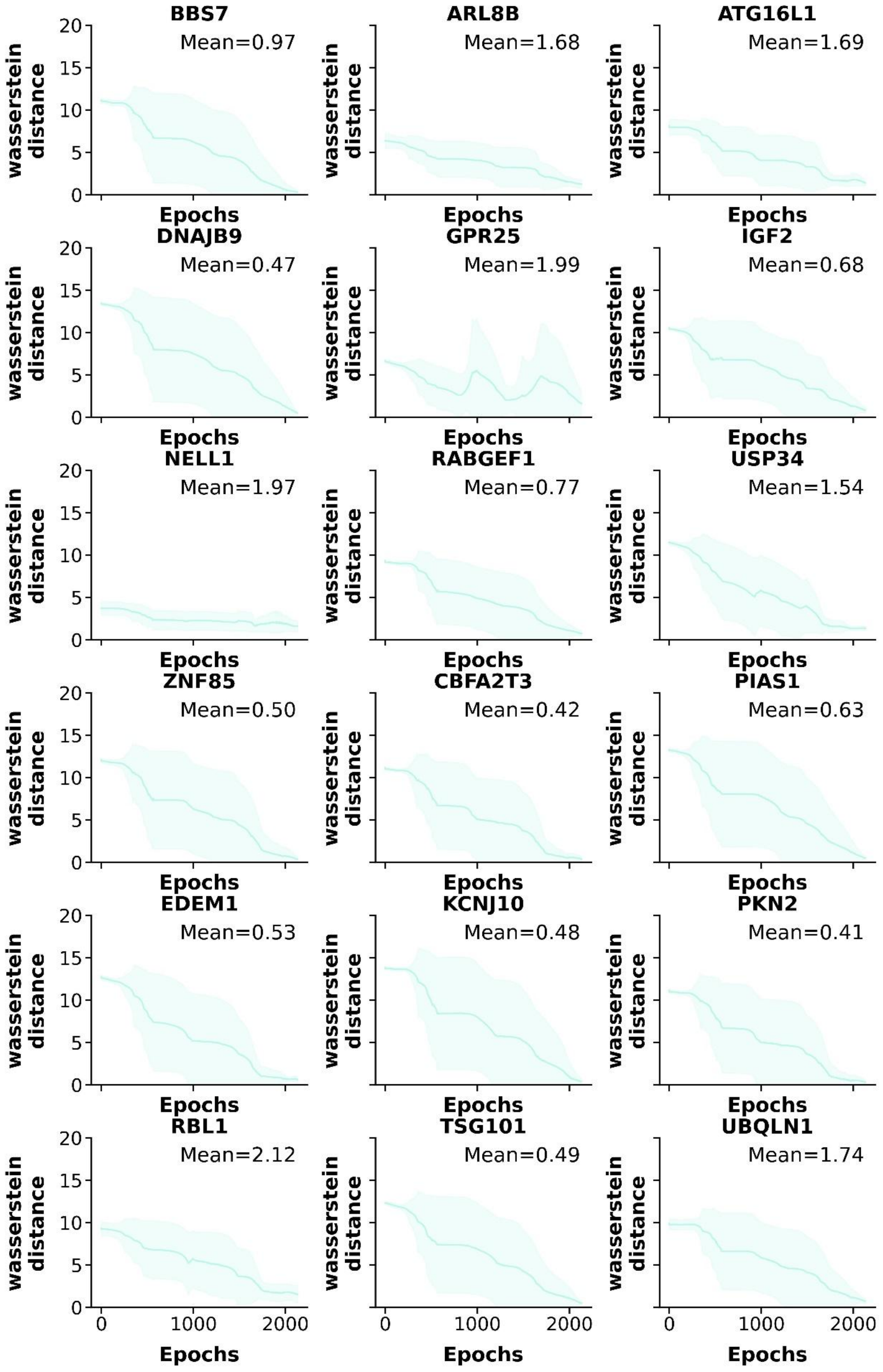


**Figure S4. ExoMasso's training optimization by per-mRNA Wasserstein distance.** We computed the per-gene Wasserstein distance per fold per initialized model (3 models, 3 folds) between NMIBC and uEV during ExoMasso's K-fold cross validation to evaluate if ExoMasso has completed training to map uEV mRNA signatures to NMIBC mRNA signatures.

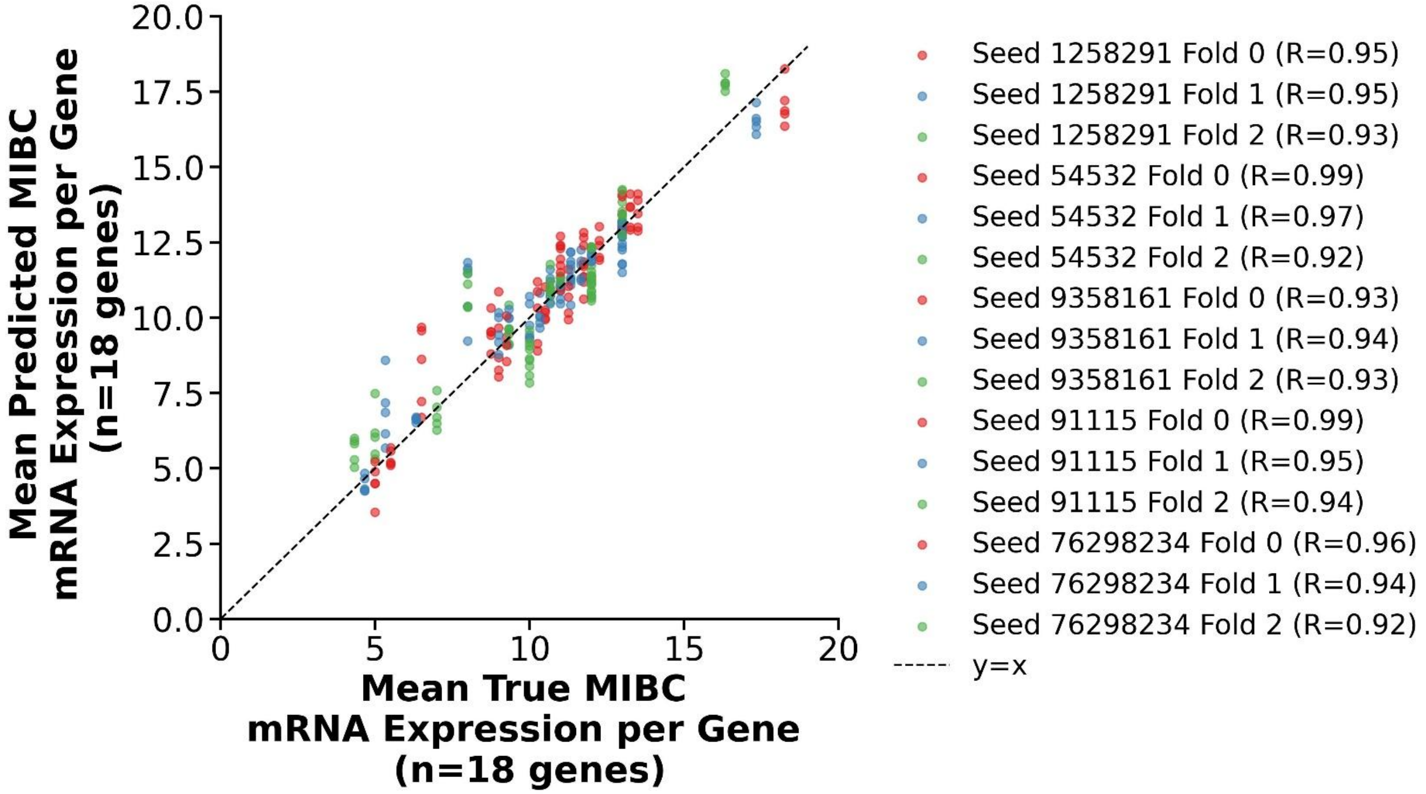


**Figure S5. ExoMasso's performance on the MIBC validation folds.** A correlation plot was computed to show the individual validation fold performance to map the mean uEV mRNA signature per fold and initialization (3 models, 3 folds) to the MIBC mRNA signature, computing the Pearson correlation coefficient (R) as a similarity measure.

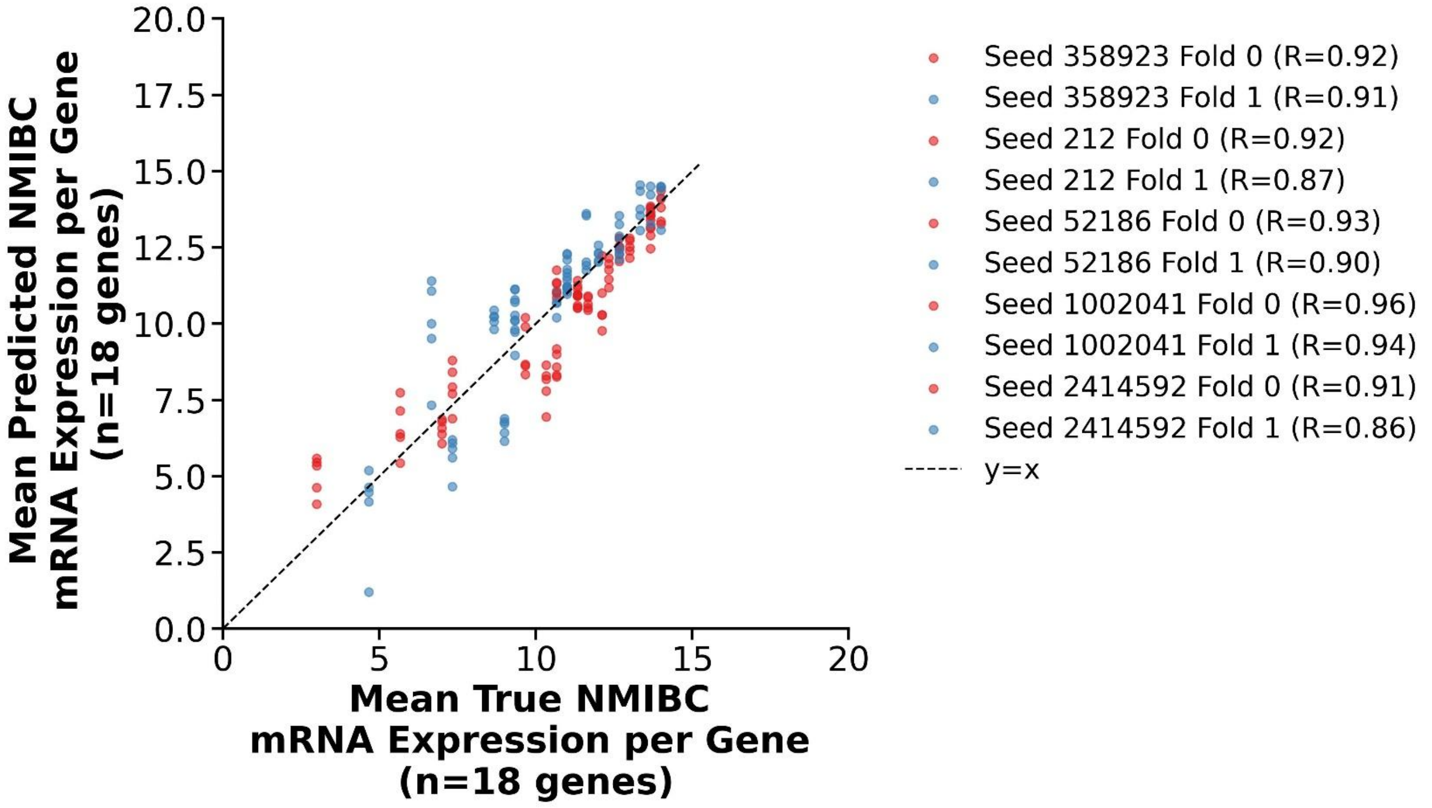


**Figure S6. ExoMasso's performance on the NMIBC validation folds.** A correlation plot was computed to show the individual validation fold performance to map the mean uEV mRNA signature per fold and initialization (3 models, 2 folds) to the NMIBC mRNA signature, computing the Pearson correlation coefficient (R) as a similarity measure.
